# The EAT-Lancet Planetary Health Diet: Impact on Cardiovascular Disease and the Environment in the EPIC Cohort

**DOI:** 10.1101/2024.06.17.24309022

**Authors:** Chiara Colizzi, Joline WJ Beulens, Reina E Vellinga, Krasimira Aleksandrova, Christina C Dahm, Inge Huybrechts, Timothy J Key, Jessica E Laine, Keren Papier, Paolo Vineis, Elisabete Weiderpass, Claudia Agnoli, Jeroen Berden, Paolo Chiodini, Jytte Halkjer, Alicia Heath, Verena Katzke, Giovanna Masala, Olatz Mokoroa, Conchi Moreno-Iribas, Genevieve Nicolas, Daniele Rodriguez-Palacios, Carlotta Sacerdote, Maria-Jose Sanchez, Matthias B. Schulze, Anne Tjønneland, W.M.Monique Verschuren, Yvonne T van der Schouw

## Abstract

**Background:** Diet plays an important role in the development of cardiovascular diseases and in maintaining sustainable planetary boundaries. The EAT-Lancet Planetary Health Diet could potentially provide co-benefits for human and environmental health, yet evidence on the association between adherence to the EAT-Lancet Planetary Health Diet and risk of cardiovascular events and environmental impact is limited.

**Methods:** We investigated the association between adherence to the EAT-Lancet diet and coronary heart disease (CHD) and stroke risk, and with greenhouse gas (GHG) emissions, land use, and dietary species richness (DSR). We included 364,745 adult men and women participating in the European Prospective Investigation into Cancer and Nutrition (EPIC) study. Food frequency questionnaires were used to create a score reflecting adherence to the EAT-Lancet diet (EAT-Lancet diet-score), ranging from 0 (no adherence) to 140 (complete adherence). A (pro-) vegetarian version of the score, the EAT-Lancet dietVV-score, was also created, which rewarded low to no consumption of all animal-based foods. Cox proportional hazard regressions were used to study the association of adherence to the EAT-Lancet diet with CHD and stroke incidence. Linear regression analyzed the association with GHG emissions, land use, and DSR.

**Findings:** Over a median follow-up of 12·8 years, we identified 12,690 CHD and 7,088 stroke cases. After multivariable adjustment, those most adherent to the EAT-Lancet diet had lower risk of incident stroke (HR_Q5vsQ1_: 0·59, 95%CI = 0·54 to 0·64), and of incident CHD for those younger than 60 years old at baseline (HR_Q5vsQ1_: 0·86, 95%CI = 0·79 to 0·93). High adherence to the EAT-Lancet diet reduced GHG emissions by 1·7% (95%CI = -1·9 to -1·5) and land use by 6·2% (95%CI = -6·4 to -5·9). The EAT-Lancet dietVV-score further reduced GHG emissions and land use by 14·3% (95%CI= -14·5 to -14·0) and 18·8% (95%CI = -19·0 to -18·5), respectively, when comparing extreme quintiles, while hazard ratios for CHD and stroke remained unchanged. Those most adherent to the EAT-Lancet diet consumed 16·1% (95%CI = 15·9 to 16·4) more plant species and 19·7% (95%CI = -20·11 to -19·40) fewer animal species.

**Interpretation:** Higher adherence to the EAT-Lancet diet was associated with co-benefits for both cardiovascular outcomes and environmental indicators, including dietary species richness. Lower GHG emissions and land use were achieved by further reducing consumption of animal-based products.

**Funding:** The coordination of EPIC-Europe is financially supported by the International Agency for Research on Cancer (IARC) and also by the Department of Epidemiology and Biostatistics, School of Public Health, Imperial College London which has additional infrastructure support provided by the NIHR Imperial Biomedical Research Centre (BRC). Exposure indicators were calculated with financial support of the Wereld Kanker Onderzoek Fonds (WKOF), as part of the World Cancer Research Fund International grant programme (IIG_FULL_2020_034).

**Research in context:** *Evidence before this study:* The authors considered all evidence available to them on the EAT-Lancet Planetary Health Diet, published up until May 2024. The authors searched for relevant articles on the association between adherence to the diet and cardiovascular outcomes and environmental indicators. Studies investigating the association between the diet and outcomes not of interest in this study were not considered. We restricted to evidence from prospective cohort studies with similar analyses and methodology, thereby excluding studies modelling the environmental impact. We found two research articles that explored the association of EAT-Lancet Planetary Health Diet with both cardiovascular outcomes and environmental impact, four that only assessed the association with cardiovascular outcomes, and three only focused on environmental indicators. We found no studies on the association between adherence to the EAT-Lancet Planetary Health Diet and food biodiversity. These studies spanned across varied population groups, focused on different cardiovascular endpoints and reported inconclusive evidence. This also streams from the use of different scores and indices to measure adherence to the EAT-Lancet Planetary Health Diet, which strongly influences evidence on risk estimates. Similarly, evidence on greenhouses gas emissions and land use are hindered by the use of different methodologies to calculate the associated environmental impact of foods and beverages.

*Added value of this study:* This study benefits from the use of a large pan-European cohort, which used a standardized nutrient and food database to determine individual dietary intake, as well as environmental data derived by Life Cycle Assessment analyses validated at the European level. The use of two diet scores—one representing an omnivorous version of the EAT-Lancet Planeatry Health Diet (EAT-Lancet diet-score) and the other representing a plant-based variation (EAT-Lancet dietVV-score) —demonstrates that greater environmental benefits can be achieved with the EAT-Lancet dietVV-score by further restricting consumption of animal-based products, without impacting the benefits on human health. The study adds to the current evidence on the impact of the EAT-Lancet Planetary Health Diet on both cardiovascular health and environmental well-being, and additionally supports evidence of an association between adherence to the EAT-Lancet Planetary Health Diet and food biodiversity. The association with food biodiversity adds an important complementary measure of health and sustainability to the current body of evidence on co-benefits of the EAT-Lancet Planetary Health Diet.

*Implications of all the available evidence:* Our findings substantiate the co-benefits of adherence to the EAT-Lancet Planeatry Health Diet found in previous studies for cardiovascular health and environmental indicators, with evidence from a large pan-European population-based study. This research study found evidence that adherence to the EAT-Lancet diet was associated with lower risk of stroke across the whole population and with lower risk of CHD among those younger than 60 years old. This study also highlights the impact of the ways in which we operationalise adherence to the EAT-Lancet Planetary Health Diet, emphasizing its importance for comparing studies and developing national policies.

## Introduction

Our current food production processes put high pressure on environmental resources, with agriculture taking up more than 43% of habitable land and contributing to 26% of global greenhouse gas (GHG) emissions, resulting in profound adverse effects on biodiversity and resilience of ecosystems ^1,2^. At the same time, the United Nations estimates that the world population will continue to grow from 7·7 billion people in 2019 to 9·7 billion people in 2050 ^3^. Addressing the nutritional needs of this expanding world population while ensuring access to adequate and healthy diets is therefore becoming an increasing global concern.

In response to this challenge, the EAT-Lancet Commission on Healthy Diets From Sustainable Food Systems proposed the Planetary Health diet (EAT-Lancet diet) ^4^. The EAT-Lancet diet is a universal diet emphasizing intake of plant-based foods and suggesting a limited intake of animal-sourced foods and starchy vegetables. The EAT-Lancet diet Commission has estimated that global adoption of the EAT-Lancet diet would prevent 10·8 to 11·6 million deaths per year, equaling 19-24% of total deaths among adults. Simultaneously, adherence to the EAT-Lancet diet would aid in keeping within the earth’s food production boundaries with regard to environmental resources, although the Commission estimates that other measures, such as reducing food waste and improving production practices, are needed as well ^4^.

The health and environmental effects have been modelled by the EAT-Lancet diet Commission, yet empirical evidence from population-based cohort studies is still scarce. Both coronary heart disease (CHD) and stroke are important drivers of mortality, accounting for 5·3 million and 5·5 million annual deaths, respectively, and dietary factors have been attributed to a higher risk of both diseases. ^5,6^. Five studies have explored the association of the EAT-Lancet diet with risk of CHD and stroke, showing inconsistent findings ^7–11^, which may be partly due to the heterogenous use of scoring systems ^12,13^. Because the EAT-Lancet proposed diet is meant to be healthy and sustainable, it is also essential to assess the environmental impact of the diet, by using two gold-standard indicators of sustainability, GHG emissions and land use. However, current evidence on the environmental impact of the EAT-Lancet diet remains fragmented and dependent on the methodology used to assess environmental impact ^9,14,15^.

Additionally, increasing food biodiversity is a strategy that co-benefits human nutrition and environmental well-being. Indeed, studies have found that increasing diversity in someone’s diet leads to a greater probability of eating a wide range of (potentially) nutritious and healthy foods ^16–18^. At the same time, increasing food biodiversity helps minimize the risk of ecosystems being disrupted by overconsumption of one single species ^17–19^. Dietary species richness (DSR) is the sum of the number of species consumed per day on average, and is a measure of food biodiversity ^20^. Thus, assessing the species richness associated with the EAT-Lancet diet provides us with the oppurtunity to add a complementary dimension in the association of the EAT-Lancet diet with human and environmental health that has not been previously explored. Therefore, this research aims at studying the adherence to the EAT-Lancet diet in a pan-European study and its association with cardiovascular events, indicators of environmental impact, and food biodiversity.

## Methods

### Study population

We used data from the European Prospective Investigation into Cancer and Nutrition (EPIC-Europe) study: an ongoing prospective cohort study of 521,323 men and women aged between 25 to 70 years at baseline between 1992 and 2000 from 23 centers across 10 European countries. For the present analysis we used data from Italy, Spain, the United Kingdom (UK), the Netherlands, Germany, Denmark, and Sweden, and excluded participants from Greece (n=28,561) and Norway (n=36,442) due to an unresolved data protection regulation issue and participants from France due to inconsistencies in the outcome definition with the other EPIC-centers (n=74,523). We further excluded participants with missing diet assessment data (n=6,310), and those with implausible energy intakes defined as being in the top and bottom 1% of the distribution of the ratio of energy intake over estimated energy requirement (n=8,196). Additionally excluding participants with prevalent CHD or stroke at baseline (n=2,546) resulted in an analytical sample of 364,745 participants **(Supplemental Figure 1).**

Most centers recruited participants from the general population, with a few exceptions. First, participants from the centers in Utrecht (Netherlands) and Florence (Italy) were recruited through a population-based breast cancer screening program. Second, participants from some of the Spanish and Italian centers were recruited from local blood donor associations. In Oxford (UK), half of the cohort was recruited among (lacto-ovo) vegetarian and vegan individuals, thereby representing a generally ‘health-conscious’ cohort. The cohorts in Utrecht (Netherlands) and Naples (Italy) only included women. Detailed information on the rationale and design of EPIC has been described previously ^21,22^. The EPIC study was approved by the Ethical Review Boards of the International Agency for Research on Cancer (IARC) and the Institutional Review Board of each participating EPIC center.

### Diet assessment

At baseline, habitual dietary intake over the past 12 months was assessed using country-specific validated dietary questionnaires. Most centers used a validated (semi-)quantitative food frequency questionnaire (FFQ), although a combination of dietary assessment methods was used in Malmö (Sweden). Nutrient and food intakes were derived through the standardized EPIC Nutrient Database, which contains over 11,000 food and beverage items ^23^.

Dietary intake assessments were used to calculate adherence to the EAT-Lancet diet. The EAT-Lancet diet score was constructed as described by Colizzi et al. ^9^. This proportional scoring method has been found to reflect well adherence to the EAT-Lancet diet recommendations and was preferred over binary-style scoring methods in a recent systematic review ^24^. Participants were assigned a proportional score ranging between 0-10 points for each dietary recommendation in the EAT-Lancet diet, totaling to a score between 0 (no adherence) and 140 points (complete adherence). The methodology of the scoring was informed by the Dutch Healthy Diet 2015 index distinguishing adequacy, moderation, optimum and ratio components **(Supplemental Figure 2)** ^25^. The dietary recommendations for the food groups in the EAT-Lancet diet are based on intakes of 2500 kcal per day for both men and women ^4^. As energy requirements for women are lower, we re-calculated the food group recommendations to 2000 kcal/day for women (except for the ratio-component and fiber intake, since we deemed these to be energy-independent).

The scoring approach is shown in **Supplemental Table 1** and described in detail elsewhere ^9^. In brief, all recommendations were scored proportionally from 0-10 as an adequacy (whole grains, fruit, vegetables, non-soy legumes, soy foods), optimum (dairy, starchy vegetables, chicken and other poultry, eggs, and fish), moderation (red and processed meat, sweets), or ratio component (unsaturated and saturated fats) ^9^. For this study, two sub-components were created for the recommendation on whole grains (including rice, wheat, corn and other grains). As information on type of cereal (e.g., wholegrain) was not available, fiber intake was used as an indicator of wholegrain consumption. This sub-component was scored as an adequacy component, with participants being assigned 5 points if they had an intake equaling or larger than 30g of fiber per day, 0 points for no consumption, and a proportional score for intermediate intakes. The second subcomponent reflected the recommendation to limit consumption of (dry) grains to 232g/day, equaling to 464g in converted wet weight ^26^, for which participants were assigned 5 points or 0 points, for meeting or not meeting the recommendation, respectively.

In the original scoring approach, consumption of selected animal-sourced foods (whole milk or derivative equivalents, chicken and other poultry, eggs, and fish) was scored as optimum components and assigned zero points or a proportionally increasing score, respectively for no or low intake and 10 points for the optimum intake ^9^. In order to reflect adherence among those eating (pro) vegetarian and vegan diets, we also constructed an alternative score, for which dairy, chicken, eggs, and fish were scored as moderation components, meaning those participants with no or low intakes of animal-sourced foods were assigned 10 points. We refer to this alternative score as the EAT-Lancet dietVV-score.

### Cardiovascular events ascertainment

Incident CHD was defined as any first fatal or non-fatal CHD event, which was a composite myocardial infarction (International Classification of Diseases, 10th Revision (ICD-10) codes I21, I22), angina (ICD-10 code I20) and other types of acute or chronic ischemic heart diseases (ICD-10 codes I23, I24, I25). Incident stroke was defined as any first fatal or non-fatal stroke event, which was a composite of hemorrhagic stroke (I60-I61), ischemic stroke (I63), unclassified stroke (I64), and other acute cerebrovascular events (I62, I65-69, F01). Fatal outcome events were generally ascertained through linkage with death registries. Non-fatal outcome events were ascertained through a variety of methods across centers, including follow-up questionnaires or linkage with morbidity/hospital registries.

### Environmental impact assessment

Environmental impact of adhering to the EAT-Lancet diet was measured via two indicators, GHG emissions and land use. The GHG emissions and land use associated with each food and drink measured via the dietary intake assessments were calculated using SHARP Indicator Database (SHARP-ID). More details on this database can be found elsewhere^27^. Briefly, SHARP-ID uses life cycle analyses to calculate the GHG emissions and land use of 994 food items coded with a unique FoodEx2-code from the FoodEx2 Exposure Hierarchy of the European Food Safety Authority (EFSA)^28^. Items included in the SHARP-ID are based on the reported food intake of four European countries included in the SUSFANS project (2005-2008), i.e. Denmark, Czech Republic, Italy and France. The database’s system boundaries includes cradle to plate, excluding industrial food processing, storage, and transport from local retailers to home. SHARP-ID applies economical allocation (based on economic value) for all non-animal-sources food, whereas nitrogen content is used for animal-source foods. For the present analysis, the environmental impact of each food group included in the EAT-Lancet diet score was summed into two variables describing the GHG emissions and land use of the diet.

### Food Biodiversity assessment

In this study we use DSR as indicator of food biodiversity. DSR was measured as described by Lachat et al. ^20^ and Hanley-Cook et al. ^17^. Shortly, DSR is calculated by the sum of the number of all species (both plant- and animal-based) consumed by each individual. For this cohort, DSR was calculated using all unique biological species of all foods and drinks using the EFSA’s Food-Ex2 food classification system ^28^. Composite dishes were decomposed into single ingredients and then into species. All food items eaten “never or less than once per month” were not included in the DSR. A total of 108 species were included in the data. Quantities were not considered when calculating the DSR, as irrelevant towards the overall count of species consumed ^17^. The DSR results in a total count of species consumed, expressed continuously. Additionally, species were split for further analyses into DSRPlant and DSRAnimal, representing respectively the total number of plant-based species and the total amount of animal-based species. All food items that could not be uniquely identified with either a plant or animal species were excluded from the total. These included food items such as mixed fats, confectionary and cakes, alcoholic and non-alcoholic beverages, condiments and dressings.

### Assessment of other covariates

Data on socio-demographic (age, sex), lifestyle and other factors were collected at baseline through validated questionnaires ^21^. Educational level was categorized into primary school, technical/professional school, secondary school, and longer education (including university degree). Alcohol intake was a continuous variable, measured in grams per day. Physical activity was assessed through the Cambridge Physical Activity index, which captures occupational physical activity and other physical exercise (e.g., cycling, walking), and was used to categorize participants as inactive, moderately inactive, moderately active, and active. Smoking status was categorized into never, former, and current smoking.

Body mass index (BMI) was calculated from measured height and weight, although for participants from Norway and for some participants from France and the UK these data were self-reported. BMI was categorized into healthy weight (<25 kg/m^2^), overweight ( ≥25 - <30 kg/m^2^), and obese (≥30 kg/m^2^). Waist circumference was measured either at the narrowest circumference of the torso or at the midpoint between the lower ribs and the iliac crest. Hypertension, hyperlipidemia, and diabetes status were self-reported.

### Statistical analysis

Statistical analyses were conducted in R 4.2.3 and the α threshold for significance was set at P<0·05. Baseline characteristics were presented across quintiles of EAT-Lancet diet-adherence and for the full cohort. Normally distributed continuous variables were presented as means with standard deviations, and skewed continuous variables were presented as medians and interquartile range (IQR). Categorical variables were presented as counts and percentages. For some of these covariates there were missing data (educational level n=15,317; physical activity n=7,220; smoking status n=2,999; waist circumference n=61,571; hypertension n=71,536; hyperlipidemia n=109,366; diabetes status n=34,648). These missings were imputed using multiple imputation methods ^29^. We used the R package MICE ^30^, using 10 imputation sets and 10 burn-it iterations. The assumption of missing at random (MAR) was checked before conducting multiple imputation. MICE *pool() function* was used to combine results from the Cox proportional hazard regression regression analyses across the imputed datasets using Rubin’s rule ^29,30^.

Cox proportional hazard regression regression was used to evaluate the association between quintiles of adherence to the EAT-Lancet diet-score and CHD and stroke incidence, with the lowest quintile as reference. Additionally, associations were examined linearly with EAT-Lancet diet-scores modelled per 10-point increment. Confounder adjustments were determined a priori and were informed by literature. In model 1 we adjusted for age and sex; in model 2 we further adjusted for educational level, smoking, alcohol consumption, physical activity, and energy intake. In model 3 we additionally adjusted for cardiovascular risk factors, such as BMI, waist circumference, hypertension, hyperlipidemia, and diabetes status. In this study, we consider model 3 to be the main model. All models were stratified by EPIC study center. The proportional hazards assumption was tested using the Schoenfeld test, indicating no violation of the assumption. Analyses were repeated for the EAT-Lancet dietVV-score.

To examine whether associations were consistent across various subgroups, we tested for interaction for levels of age, sex, educational level, and BMI. Interaction was tested by using a likelihood ratio test (LRT) for a model with and without interaction term, for each imputed dataset. When the LRT was statistically significant (p-value <0·05) for all imputation datasets, we conducted separate analyses by subgroups. For this analysis, age was categorized in two groups, those younger than 60 years old and those older than 60. Additionally, we carried out a number of sensitivity analyses. First, we excluded the first 2 years of follow-up, as subclinical disease may have affected eating habits and have induced reverse causation. Second, we excluded food group recommendations individually in order to explore the food groups driving the association. Finally, we measured adherence to the EAT-Lancet diet across countries and centers.

We used linear regression models to estimate the association between EAT-Lancet diet-adherence and GHG emissions, land use, and DSR. GHG emissions and land use were calculated as the sum of the associated environmental impact of all the food groups included in the EAT-Lancet diet. The lowest quintile was used as reference. In model 1, we adjusted for age, sex, and energy intake, as informed by the literature ^31–33^. Bootstrapping was used to estimate the percentage difference between Q1 and Q5 and its 95% CI. Additionally, the impact on GHG emissions, land use, and DSR was estimated using the EAT-Lancet dietVV-score.

### Roles of the funding source

The funders of the study did not play a role in data collection, data analysis, data interpretation, writing of the manuscript, or the decision to submit for publication.

## Results

### Baseline characteristics

The EAT-Lancet diet-score in the population ranged from 10 to 120 and the mean was 64 (SD=15) . Participants generally scored most points on the recommendations for grains (rice, wheat and corn), added fats, and dairy, and the least points on the recommendations for red meat, added sugar, legumes, soy and nuts. Participants in Spain, UK, and Italy had the highest average EAT-Lancet diet-scores and in Sweden, the Netherlands, and Germany the lowest (**Supplemental Table 2**). Participants in the highest EAT-Lancet diet-adherence quintile were more likely to be female, non-smokers, had higher total energy intake, lower alcohol consumption, and smaller waist circumference (**Table 1**).

**Table 1.**
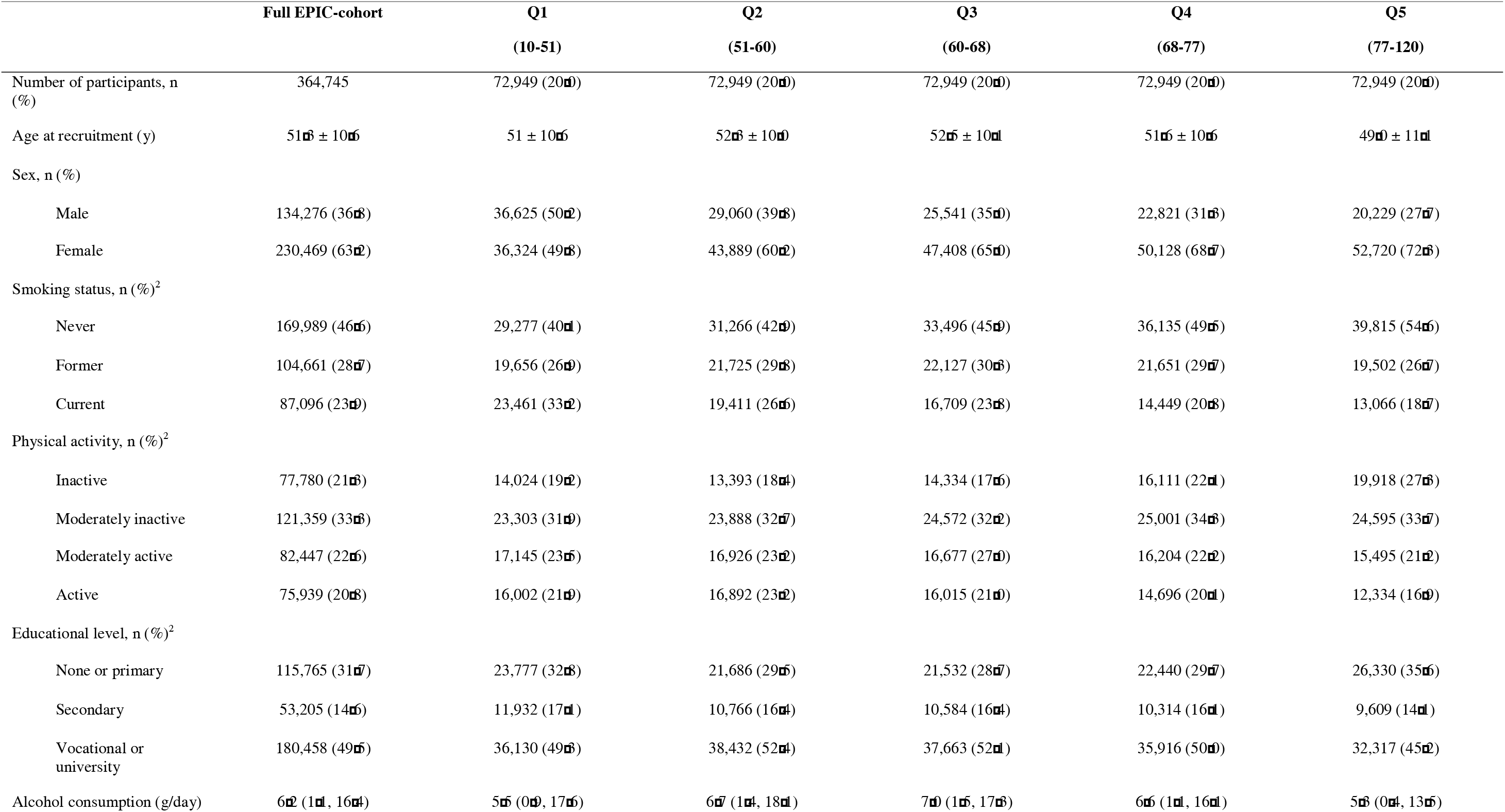
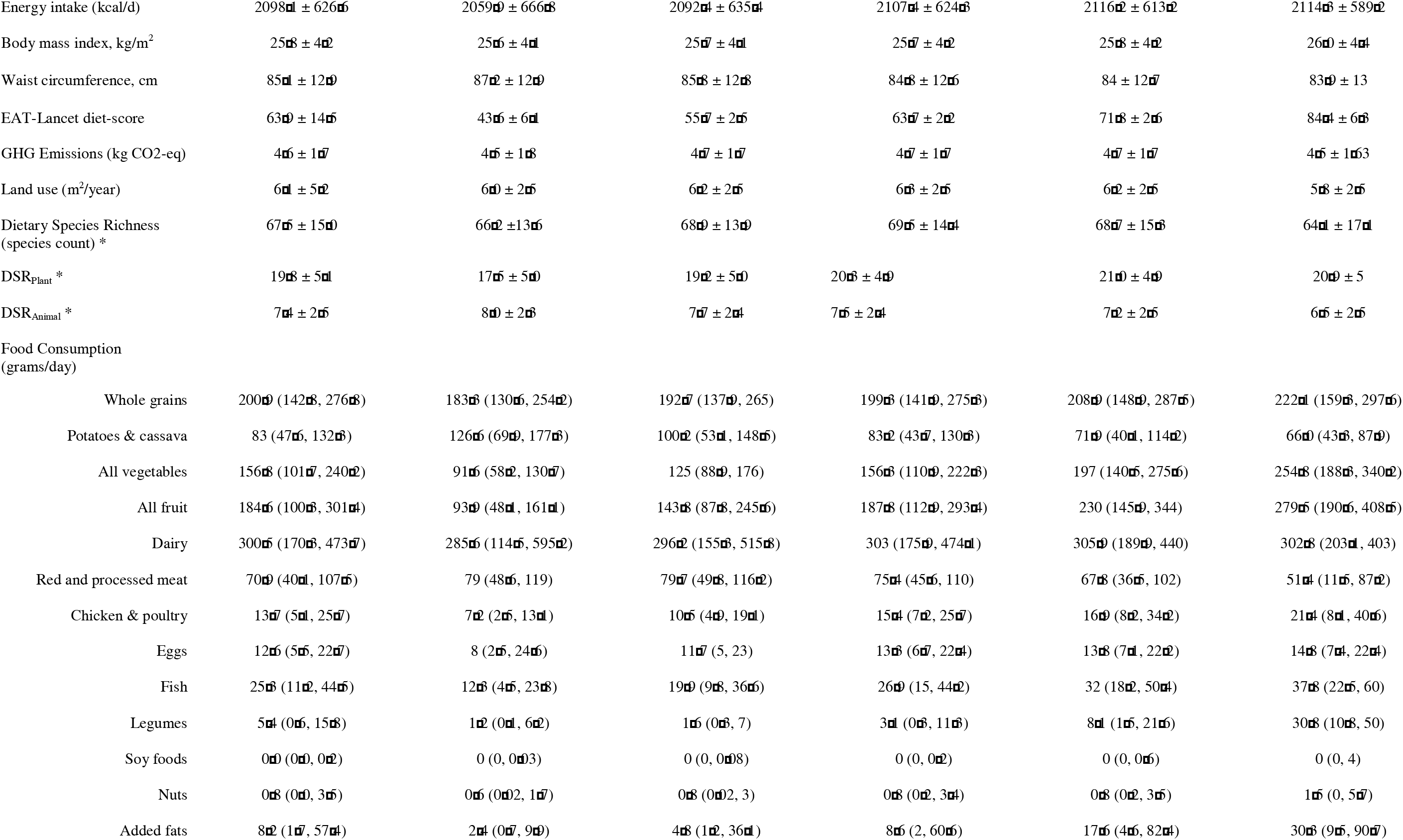
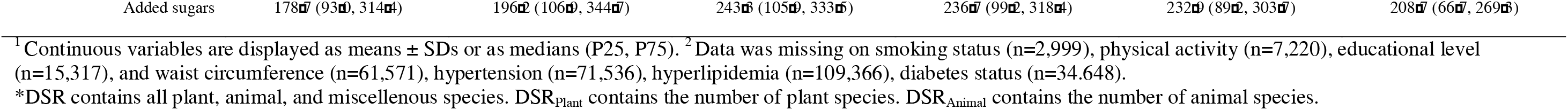
Baseline characteristics across quintiles of EAT-Lancet diet-score (N=364,745)^1^.

|  | Full EPIC-cohort | Q1<br>(10-51) | Q2<br>(51-60) | Q3<br>(60-68) | Q4<br>(68-77) | Q5<br>(77-120) |
| --- | --- | --- | --- | --- | --- | --- |
| Number of participants, n (%) | 364,745 | 72,949 (20.0) | 72,949 (20.0) | 72,949 (20.0) | 72,949 (20.0) | 72,949 (20.0) |
| Age at recruitment (y) | 51.3 ± 10.6 | 51 ± 10.6 | 52.3 ± 10.0 | 52.5 ± 10.1 | 51.6 ± 10.6 | 49.0 ± 11.1 |
| Sex, n (%) |  |  |  |  |  |  |
| Male | 134,276 (36.8) | 36,625 (50.2) | 29,060 (39.8) | 25,541 (35.0) | 22,821 (31.3) | 20,229 (27.7) |
| Female | 230,469 (63.2) | 36,324 (49.8) | 43,889 (60.2) | 47,408 (65.0) | 50,128 (68.7) | 52,720 (72.3) |
| Smoking status, n (%)<sup>2</sup> |  |  |  |  |  |  |
| Never | 169,989 (46.6) | 29,277 (40.1) | 31,266 (42.9) | 33,496 (45.9) | 36,135 (49.5) | 39,815 (54.6) |
| Former | 104,661 (28.7) | 19,656 (26.9) | 21,725 (29.8) | 22,127 (30.3) | 21,651 (29.7) | 19,502 (26.7) |
| Current | 87,096 (23.9) | 23,461 (33.2) | 19,411 (26.6) | 16,709 (23.8) | 14,449 (20.8) | 13,066 (18.7) |
| Physical activity, n (%)<sup>2</sup> |  |  |  |  |  |  |
| Inactive | 77,780 (21.3) | 14,024 (19.2) | 13,393 (18.4) | 14,334 (17.6) | 16,111 (22.1) | 19,918 (27.3) |
| Moderately inactive | 121,359 (33.3) | 23,303 (31.9) | 23,888 (32.7) | 24,572 (32.2) | 25,001 (34.3) | 24,595 (33.7) |
| Moderately active | 82,447 (22.6) | 17,145 (23.5) | 16,926 (23.2) | 16,677 (27.0) | 16,204 (22.2) | 15,495 (21.2) |
| Active | 75,939 (20.8) | 16,002 (21.9) | 16,892 (23.2) | 16,015 (21.0) | 14,696 (20.1) | 12,334 (16.9) |
| Educational level, n (%)<sup>2</sup> |  |  |  |  |  |  |
| None or primary | 115,765 (31.7) | 23,777 (32.8) | 21,686 (29.5) | 21,532 (28.7) | 22,440 (29.7) | 26,330 (35.6) |
| Secondary | 53,205 (14.6) | 11,932 (17.1) | 10,766 (16.4) | 10,584 (16.4) | 10,314 (16.1) | 9,609 (14.1) |
| Vocational or university | 180,458 (49.5) | 36,130 (49.3) | 38,432 (52.4) | 37,663 (52.1) | 35,916 (50.0) | 32,317 (45.2) |
| Alcohol consumption (g/day) | 6.2 (1.1, 16.4) | 5.5 (0.9, 17.6) | 6.7 (1.4, 18.1) | 7.0 (1.5, 17.3) | 6.6 (1.1, 16.1) | 5.3 (0.4, 13.5) |
| Energy intake (kcal/d) | 2098.1 ± 626.6 | 2059.9 ± 666.8 | 2092.4 ± 635.4 | 2107.4 ± 624.3 | 2116.2 ± 613.2 | 2114.3 ± 589.2 |
| Body mass index, kg/m<sup>2</sup> | 25.8 ± 4.2 | 25.6 ± 4.1 | 25.7 ± 4.1 | 25.7 ± 4.2 | 25.8 ± 4.2 | 26.0 ± 4.4 |
| Waist circumference, cm | 85.1 ± 12.9 | 87.2 ± 12.9 | 85.8 ± 12.8 | 84.8 ± 12.6 | 84 ± 12.7 | 83.9 ± 13 |
| EAT-Lancet diet-score | 63.9 ± 14.5 | 43.6 ± 6.1 | 55.7 ± 2.5 | 63.7 ± 2.2 | 71.8 ± 2.6 | 84.4 ± 6.3 |
| GHG Emissions (kg CO2-eq) | 4.6 ± 1.7 | 4.5 ± 1.8 | 4.7 ± 1.7 | 4.7 ± 1.7 | 4.7 ± 1.7 | 4.5 ± 1.63 |
| Land use (m<sup>2</sup>/year) | 6.1 ± 5.2 | 6.0 ± 2.5 | 6.2 ± 2.5 | 6.3 ± 2.5 | 6.2 ± 2.5 | 5.8 ± 2.5 |
| Dietary Species Richness<br>(species count) * | 67.5 ± 15.0 | 66.2 ± 13.6 | 68.9 ± 13.9 | 69.5 ± 14.4 | 68.7 ± 15.3 | 64.1 ± 17.1 |
| DSR<sub>plant</sub> * | 19.8 ± 5.1 | 17.5 ± 5.0 | 19.2 ± 5.0 | 20.3 ± 4.9 | 21.0 ± 4.9 | 20.9 ± 5 |
| DSR<sub>Animal</sub> * | 7.4 ± 2.5 | 8.0 ± 2.3 | 7.7 ± 2.4 | 7.5 ± 2.4 | 7.2 ± 2.5 | 6.5 ± 2.5 |
| Food Consumption<br>(grams/day) |  |  |  |  |  |  |
| Whole grains | 200.9 (142.8, 276.8) | 183.3 (130.6, 254.2) | 192.7 (137.9, 265) | 199.3 (141.9, 275.3) | 208.9 (148.9, 287.5) | 222.1 (159.3, 297.6) |
| Potatoes & cassava | 83 (47.6, 132.3) | 126.6 (69.9, 177.3) | 100.2 (53.1, 148.5) | 83.2 (43.7, 130.3) | 71.9 (40.1, 114.2) | 66.0 (43.3, 87.9) |
| All vegetables | 156.8 (101.7, 240.2) | 91.6 (58.2, 130.7) | 125 (88.9, 176) | 156.3 (110.9, 222.3) | 197 (140.5, 275.6) | 254.8 (188.3, 340.2) |
| All fruit | 184.6 (100.3, 301.4) | 93.9 (48.1, 161.1) | 143.8 (87.8, 245.6) | 187.8 (112.9, 293.4) | 230 (145.9, 344) | 279.5 (190.6, 408.5) |
| Dairy | 300.5 (170.3, 473.7) | 285.6 (114.5, 595.2) | 296.2 (155.3, 515.8) | 303 (175.9, 474.1) | 305.9 (189.9, 440) | 302.8 (203.1, 403) |
| Red and processed meat | 70.9 (40.1, 107.5) | 79 (48.6, 119) | 79.7 (49.8, 116.2) | 75.4 (45.6, 110) | 67.8 (36.5, 102) | 51.4 (11.5, 87.2) |
| Chicken & poultry | 13.7 (5.1, 25.7) | 7.2 (2.5, 13.1) | 10.5 (4.9, 19.1) | 15.4 (7.2, 25.7) | 16.9 (8.2, 34.2) | 21.4 (8.1, 40.6) |
| Eggs | 12.6 (5.5, 22.7) | 8 (2.5, 24.6) | 11.7 (5, 23) | 13.3 (6.7, 22.4) | 13.8 (7.1, 22.2) | 14.8 (7.4, 22.4) |
| Fish | 25.3 (11.2, 44.5) | 12.3 (4.5, 23.8) | 19.9 (9.8, 36.6) | 26.9 (15, 44.2) | 32 (18.2, 50.4) | 37.8 (22.5, 60) |
| Legumes | 5.4 (0.6, 15.8) | 1.2 (0.1, 6.2) | 1.6 (0.3, 7) | 3.1 (0.3, 11.3) | 8.1 (1.5, 21.6) | 30.8 (10.8, 50) |
| Soy foods | 0.0 (0.0, 0.2) | 0 (0, 0.03) | 0 (0, 0.08) | 0 (0, 0.2) | 0 (0, 0.6) | 0 (0, 4) |
| Nuts | 0.8 (0.0, 3.5) | 0.6 (0.02, 1.7) | 0.8 (0.02, 3) | 0.8 (0.2, 3.4) | 0.8 (0.2, 3.5) | 1.5 (0, 5.7) |
| Added fats | 8.2 (1.7, 57.4) | 2.4 (0.7, 9.9) | 4.8 (1.2, 36.1) | 8.6 (2, 60.6) | 17.6 (4.6, 82.4) | 30.3 (9.5, 90.7) |
| Added sugars | 178.7 (93.0, 314.4) | 196.2 (106.9, 344.7) | 243.3 (105.9, 333.5) | 236.7 (99.2, 318.4) | 232.9 (89.2, 303.7) | 208.7 (66.7, 269.3) |

<sup>1</sup> Continuous variables are displayed as means ± SDs or as medians (P25, P75). <sup>2</sup> Data was missing on smoking status (n=2,999), physical activity (n=7,220), educational level (n=15,317), and waist circumference (n=61,571), hypertension (n=71,536), hyperlipidemia (n=109,366), diabetes status (n=34,648).
\*DSR contains all plant, animal, and miscellenous species. DSR<sub>Plant</sub> contains the number of plant species. DSR<sub>Animal</sub> contains the number of animal species.

### Association of EAT-Lancet diet-adherence with incident CHD

After a median follow-up time of 12·8 years 12,690 cases of CHD occurred. High adherence to the EAT-Lancet diet was not associated with a lower risk of incident CHD after adjustment for cardiovascular risk factors (HR_Q5vsQ1_: 0·98, 95%CI = 0·93 – 1·04; HR_10-point increment_: 0·99, 95%CI = 0·98 – 1·01) (**Table 2**). However, in subgroup analysis (p-values for interaction <0·001), those younger than 60 years old did show a modestly lower risk for CHD (HR_Q5vsQ1_: 0·86, 95%CI = 0·79 – 0·93; HR_10-point increment_: 0·96, 95% CI = 0·94 – 0·98), while those older than 60 had a higher risk of CHD (HR_Q5vsQ1_: 1·16, 95%CI = 1·06 – 1·26; HR_10-point increment_: 1·03, 95%CI = 1·01 – 1·05) **(Supplemental Figure 3)**. There were no differences in subgroups by sex, educational level, or BMI. When exploring the association of the EAT-Lancet dietVV-score with CHD risk, the association was comparable to the original EAT-Lancet score, which still includes animal products scored with an optimum component (**Table 3).** Neither exclusion of cases in the first two years of follow-up (**Supplemental Table 3**) nor the exclusion of each dietary score component from the EAT-Lancet score (**Supplemental Table 4)** altered our findings.

**Table 2.** Hazard ratio’s and 95% confidence intervals for quintiles of EAT-Lancet diet adherence with incident CHD and stroke.

| Quintiles of EAT-Lancet diet-score |  |  |  |  |  |  |
| --- | --- | --- | --- | --- | --- | --- |
|  | Q1 (10-51) | Q2 (51-60) | Q3 (60-68) | Q4 (68-77) | Q5 (77-120) | Per 10-point increment |
| CHD |  |  |  |  |  |  |
| Events, n | 2,852 | 2,745 | 2,640 | 2,355 | 2,098 | 12,690 |
| Person-years | 19,941 | 19,770 | 18,993 | 16,925 | 15,386 | 91,015 |
| Crude model | 1.00 | 0.95 (0.91 – 1.01) | 0.91 (0.87 – 0.96) | 0.81 (0.77 – 0.86) | 0.70 (0.67 – 0.74) | 0.92 (0.91 – 0.93) |
| Model 1<sup>a</sup> | 1.00 | 0.94 (0.89 – 0.99) | 0.92 (0.87 – 0.97) | 0.87 (0.83 – 0.92) | 0.92 (0.87 – 0.97) | 0.97 (0.96 – 0.99) |
| Model 2<sup>b</sup> | 1.00 | 0.98 (0.93 – 1.04) | 0.98 (0.93 – 1.04) | 0.96 (0.90 – 1.01) | 0.99 (0.94 – 1.05) | 0.99 (0.98 – 1.01) |
| Model 3<sup>c</sup> | 1.00 | 0.97 (0.92 – 1.02) | 0.97 (0.92 – 1.03) | 0.95 (0.90 – 1.01) | 0.98 (0.93 – 1.04) | 0.99 (0.98 – 1.01) |
| Stroke |  |  |  |  |  |  |
| Events, n | 1,829 | 1,674 | 1,448 | 1,231 | 906 | 7,088 |
| Person-years | 13,668 | 12,682 | 11,386 | 9,767 | 7,895 | 55,397 |
| Crude model | 1.00 | 0.92 (0.86 – 0.99) | 0.80 (0.74 – 0.85) | 0.67 (0.62 – 0.72) | 0.45 (0.42 – 0.49) | 0.84 (0.82 – 0.85) |
| Model 1<sup>a</sup> | 1.00 | 0.86 (0.80 – 0.92) | 0.74 (0.69 – 0.79) | 0.66 (0.61 – 0.71) | 0.54 (0.50 – 0.59) | 0.86 (0.84 – 0.87) |
| Model 2<sup>b</sup> | 1.00 | 0.89 (0.83 – 0.95) | 0.78 (0.73 – 0.83) | 0.71 (0.66 – 0.76) | 0.58 (0.53 – 0.63) | 0.87 (0.86 – 0.89) |
| Model 3<sup>c</sup> | 1.00 | 0.89 (0.83 – 0.95) | 0.78 (0.73 – 0.84) | 0.72 (0.67 – 0.77) | 0.59 (0.54 – 0.64) | 0.88 (0.86 – 0.89) |

<sup>a</sup> Model 1 was adjusted for age, sex; <sup>b</sup> Model 2 was adjusted for age, sex, educational level, smoking, alcohol consumption, physical activity, and energy intake; <sup>c</sup> Model 3 was additionally adjusted for BMI, waist circumference, hypertension, hyperlipidemia, and diabetes status. Models were stratified by EPIC center.

**Table 3.** Hazard ratio’s and 95% confidence intervals for quintiles of EAT-Lancet dietVV-score with incident CHD and stroke.

|  | Quintiles of EAT-Lancet dietVV-score<sup>1</sup> |  |  |  |  | Per 10-point increment |
| --- | --- | --- | --- | --- | --- | --- |
|  | Q1 (17-65) | Q2 (65-72) | Q3 (72-79) | Q4 (79-87) | Q5 (87-134) |  |
| **CHD** |  |  |  |  |  |  |
| Events, n | 3,086 | 2,755 | 2,532 | 2,486 | 1,831 | 12,690 |
| Person-years | 22,074 | 19,574 | 18,220 | 17,933 | 13,214 | 91,015 |
| Crude model | 1.00 | 0.90 (0.85 – 0.94) | 0.83 (0.78 – 0.87) | 0.81 (0.77 – 0.86) | 0.59 (0.55 – 0.62) | 0.87 (0.86 – 0.88) |
| Model 1<sup>a</sup> | 1.00 | 0.95 (0.91 – 1.00) | 0.93 (0.88 – 0.98) | 0.97 (0.92 – 1.03) | 0.88 (0.83 – 0.94) | 0.97 (0.95 – 0.98) |
| Model 2<sup>b</sup> | 1.00 | 1.00 (0.95 – 1.05) | 0.99 (0.94 – 1.04) | 1.04 (0.98 – 1.10) | 0.95 (0.89 – 1.01) | 0.99 (0.97 – 1.00) |
| Model 3<sup>c</sup> | 1.00 | 0.98 (0.93 – 1.03) | 0.97 (0.91 – 1.02) | 1.02 (0.97 – 1.08) | 0.97 (0.91 – 1.03) | 0.97 (0.91 – 1.03) |
| **Stroke** |  |  |  |  |  |  |
| Events, n | 2,038 | 1,657 | 1,432 | 1,133 | 828 | 7,088 |
| Person-years | 15,380 | 12,674 | 11,197 | 8,997 | 7,148 | 55,397 |
| Crude model | 1.00 | 0.83 (0.78 – 0.88) | 0.71 (0.67 – 0.76) | 0.56 (0.52 – 0.60) | 0.39 (0.36 – 0.42) | 0.78 (0.77 – 0.79) |
| Model 1<sup>a</sup> | 1.00 | 0.85 (0.80 – 0.91) | 0.76 (0.71 – 0.81) | 0.63 (0.58 – 0.68) | 0.54 (0.50 – 0.58) | 0.84 (0.82 – 0.86) |
| Model 2<sup>b</sup> | 1.00 | 0.90 (0.84 – 0.96) | 0.81 (0.76 – 0.87) | 0.67 (0.62 – 0.72) | 0.58 (0.54 – 0.63) | 0.86 (0.84 – 0.88) |
| Model 3<sup>c</sup> | 1.00 | 0.89 (0.83 – 0.95) | 0.80 (0.75 – 0.86) | 0.67 (0.62 – 0.72) | 0.60 (0.55 – 0.65) | 0.87 (0.85 – 0.88) |

<sup>a</sup> Model 1 was adjusted for age, sex; <sup>b</sup> Model 2 was adjusted for age, sex, educational level, smoking, alcohol consumption, physical activity, and energy intake; <sup>c</sup> Model 3 was additionally adjusted for BMI, waist circumference, hypertension, hyperlipidemia, and diabetes status. Models were stratified by EPIC center.
<sup>1</sup>No or low intakes of animal-sourced foods (e.g., those initially scored as optimum components, including whole milk or derivative equivalents, chicken and other poultry, eggs, and fish) were assigned with 10 points.

### Association of EAT-Lancet diet-adherence with incident stroke

After a median follow-up time of 12·7 years 7,088 cases of stroke occurred. High adherence to the EAT-Lancet diet was associated with a lower risk of incident stroke after adjustment for cardiovascular risk factors (HR_Q5vsQ1_: 0·59, 95%CI = 0·54 – 0·64; HR_10-point increment_: 0·88, 95%CI = 0·86 – 0·89) (**Table 2**). The association of EAT-Lancet diet-adherence with incident stroke was slightly stronger in the subgroup analysis by age (p-values for interaction <0·001), with those aged 60 years or younger (HR_Q5vsQ1_: 0·55, 95%CI = 0·49 – 0·61; HR_10-point increment_: 0·86, 95%CI = 0·84 – 0·88) having a lower risk of stroke than those older than 60 years, (HR_Q5vsQ1_: 0·65, 95%CI = 0·58 – 0·73; HR_10-point increment_: 0·90, 95%CI = 0·88 – 0·92). A stronger association was also found for males (HR_males,_ HR_Q5vsQ1_:0·54, 95%CI = 0·48 – 0·61; HR_10-point increment_: 0·85, 95%CI = 0·83 – 0·87), compared to females (HR_females,_ HR_Q5vsQ1_: 0·67, 95%CI = 0·60 – 0·76; HR_10-point increment_: 0·91, 95%CI = 0·89 – 0·93) (p-values for interaction <0·05). **(Supplemental Figure 4)**. No differences were found for subgroups of educational level and BMI.

When exploring the association of the EAT-Lancet dietVV-score with stroke risk, HRs attenuated slightly (HR_Q5vsQ1_: 0·60, 95%CI = 0·55 – 0·65; HR_10-point increment_: 0·87, 95%CI = 0·85 – 0·88) (**Table 3**). Exclusion of cases in the first two years of follow-up did not alter findings (**Supplemental Table 3**). When excluding single components of the EAT-Lancet diet, the association was attenuated to almost the null when excluding fruit (HR_10-point increment_ : 0·99, 95%CI = 0·99 – 0·99), and became slightly more inverse when excluding added fats (HR_10-point increment_ : 0·85, 95%CI = 0·84 – 0·87) (**Supplemental Table 4**).

### Association of EAT-Lancet diet-adherence with environmental indicators

The EAT-Lancet diet-related greenhouse gas emissions in the population were on average 4·62 kg CO2-eq, ranging between 0·03 to 27·55 kg CO2-eq, and land use was 6·1 m^2^ per year on average, with a range between 0·03 and 45·89 m^2^ per year. In the adjusted models, adherence to the EAT-Lancet diet was associated with lower GHG emissions (β_Q5vsQ1_ : -0·08 kg CO2-eq, 95%CI = -0·09 – -0·07), and lower land use (β_Q5vsQ1_ : -0·38 m^2^ per year, 95%CI = -0·39 – -0·36) (Table 4). This translated to 1·7% (95%CI = -1·9 – -1·5) and 6·2% (95%CI = -6·4 – -5·9) lower GHG emission and land use, respectively, between the highest and lowest quintiles of adherence.

**Table 4.**
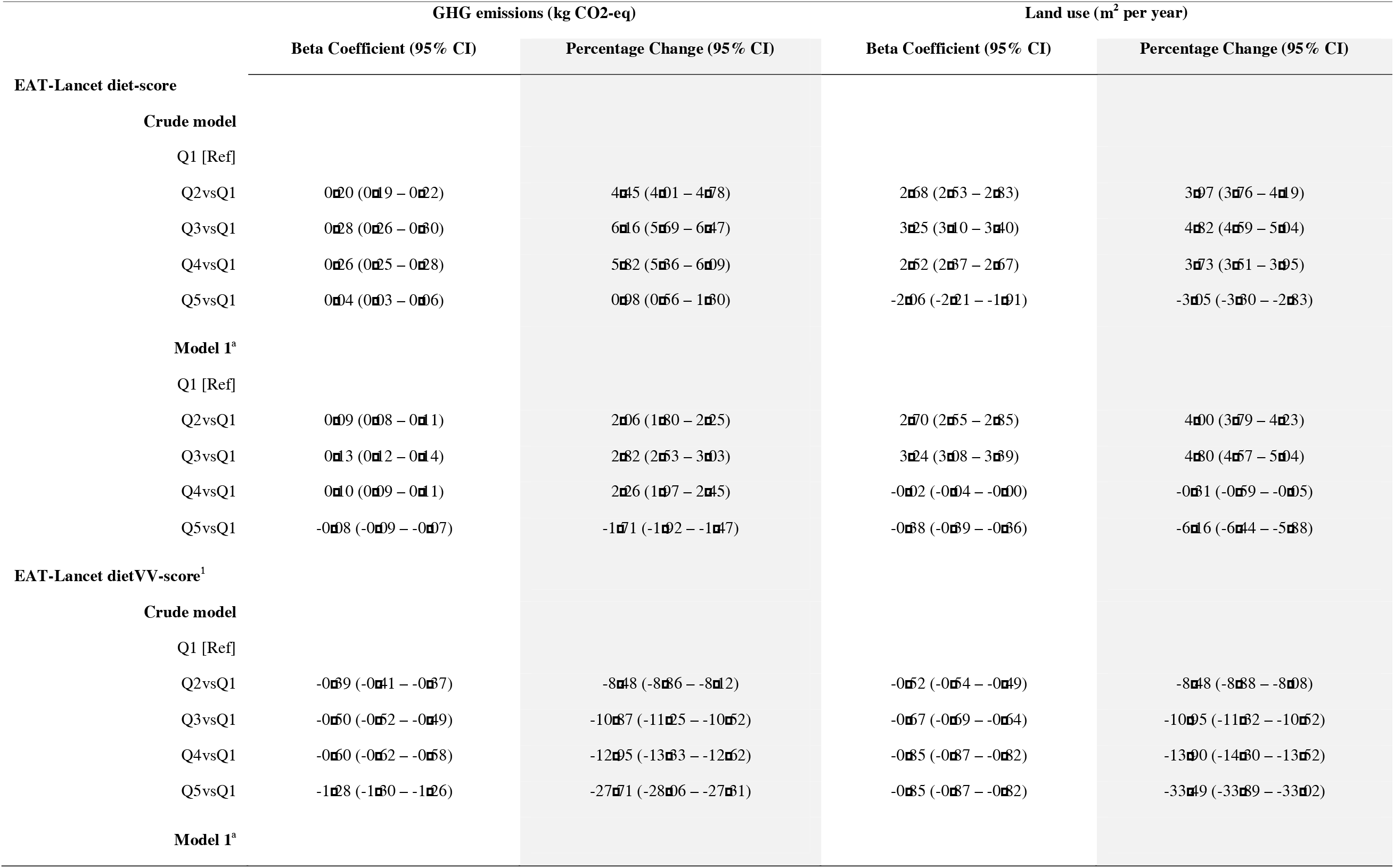
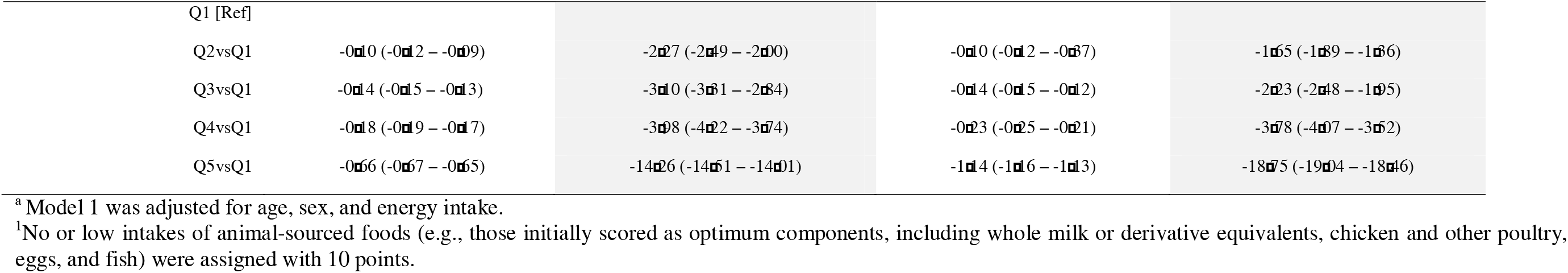
Beta coefficients, percentage change and 95% confidence intervals for quintiles of EAT-Lancet diet adherence with GHG emissions and land use.

Lower levels of both GHG emissions and land use were found when using the adjusted EAT-Lancet dietVV-score, which rewards no to very low consumption of any animal product (**Table 4)**. Further restriction of consumption of meat and animal derivates was associated with 14·3% (95%CI= -14·5 – - 14·0) less GHG emissions and 18·8% (95%CI = -19·0 – -18·5) less land use.

### Association of EAT-Lancet diet-adherence with food biodiversity

The total number of unique species count was on average equal to 67·5 (SD=15) unique species. The average total number of unique plant species was 19·8 (SD=5·1), while the mean animal species was 7·4 (SD=2·5). In the adjusted model, adherence to the EAT-Lancet diet was associated with lower DSR (β_Q5vsQ1_ : -2·43 species count, 95%CI = -2·59 – -2·28), when comparing extreme quintiles (**Table 5)**. Th translated to 3·6% (95%CI = -3·9 – -3·4) lower DSR among those with the highest adherence (Table 5). The relationship between the EAT-Lancet diet and food biodiversity differed when measuring DSR_Plant_ and DSR_Animal_ separately. In adjusted models, higher adherence to the EAT-Lancet diet was associated with higher DSR_Plant_ (β_Q5vsQ1_: 3·19 species count, 95%CI = 3·14 – 3·24) and lower DSR_Animal_ (β_Q5vsQ1_ : -1·45 species count, 95%CI = -1·50 – -1·43). When comparing extreme quintiles, this translated with adherence to the EAT-Lancet diet being associated with consuming approximately 16·1% (95%CI = 15·9 – 16·4) more plant species, and 19·7% (95%CI = -20·11 – -19·40) less animal species (**Table 5)**.

**Table 5.**
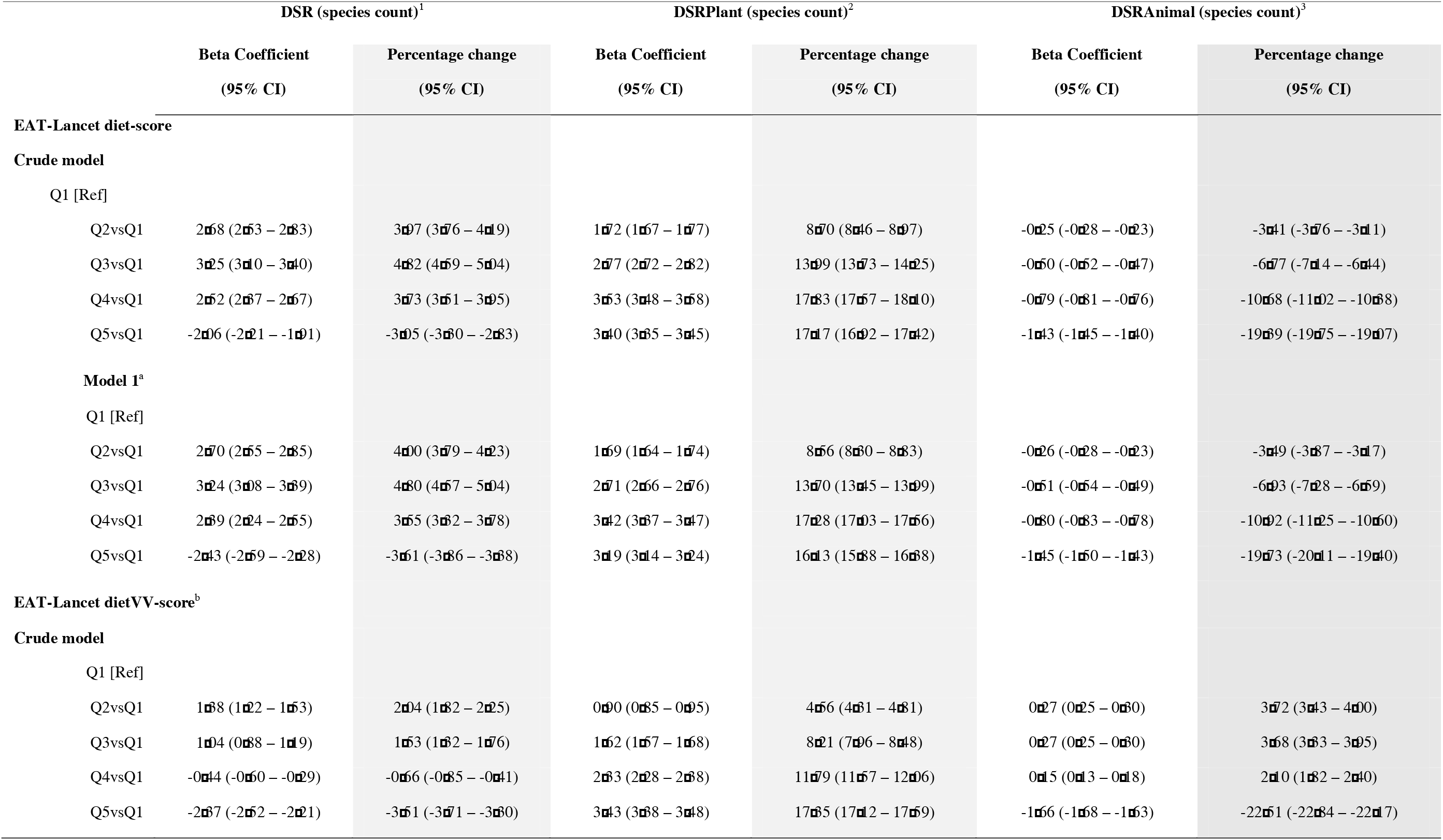
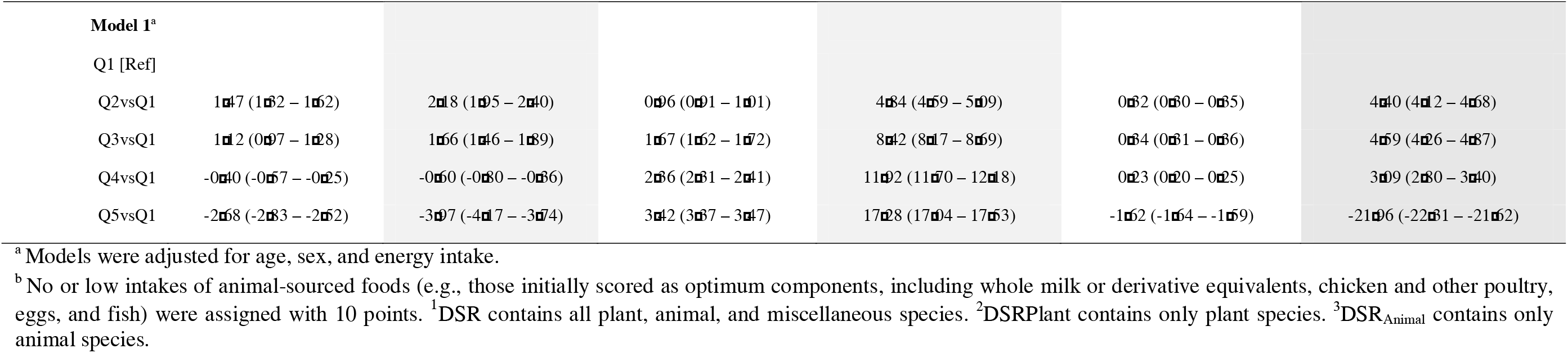
Beta coefficients, percentage change and 95% confidence intervals for quintiles of EAT-Lancet diet adherence with DSR, DSRPlant, and DSRAnimal.

Highest adherence to the (pro) vegetarian EAT-Lancet dietVV-score was associated with lower DSR than the original EAT-Lancet diet score (β_Q5vsQ1_: -2·68 species, 95%CI = -2·83 – -2·52), corresponding to 4·0% (95%CI = -4·2 – -3·7) less species consumed, when comparing extreme quintiles (**Table 5**). When considering plant and animal species separately, the EAT-Lancet dietVV-score was associated with consuming approximately 17·3% (95%CI = 17·0 – 17·5) more plant species and 22·0% (95%CI = -22·3 – -21·6) less animal species (Table 5).

## Discussion

Our study demonstrated the co-benefits of adhering to the EAT-Lancet Planetary Health Diet for cardiovascular health and environmental sustainability. We observed that higher adherence to this diet was associated with a 41% lower risk of stroke in the total population and 14% lower risk of CHD among individuals under 60 years old. Although the EAT-Lancet diet was only modestly associated with lower GHG emissions and land use, using the dietVV-score to measure adherence resulted a in 14·3% decrease in GHG emissions and an 18.8% reduction in land use, without compromising cardiovascular benefits. Additionally, adherence to the diet was associated with 16·1% more plant species richnees and 19·7% less animal species richness. In this large pan-European prospective cohort study, adherence to the EAT-Lancet diet varied between countries, with the UK, Spain, and Italy showing the highest adherence, and Sweden, the Netherlands, and Germany showing the lowest adherence. Individual recommendations for meat (e.g., lamb, beef, and pork), added sugar, soy, nuts, and legumes had particularly low scores. Generally, the maximum total EAT-Lancet diet-score was 120, indicating that in observed diets across Europe there is still room for improvement with regards to reaching the EAT-Lancet diet recommendations.

The present study is the first to report on a nuanced diet-score measuring adherence to the EAT-Lancet diet in relation to cardiovascular outcomes in a large pan-European cohort. Studies in some of the individual EPIC cohorts included in this study, namely the Dutch, Danish, Swedish, and Oxford cohorts, generally reported similar directions of associations, although statistical significance often differed (**Supplemental Tables 5 & 6**) ^7–9,11^. Potential explanations for discrepancies between this study and studies conducted in the various individual EPIC cohorts include the operationalization of the score, disease definitions, follow-up durations, and participants’ exclusions criteria. Also, dietary intake derived by the country’s specific dietary assessment method differed slightly from the dietary intake for the whole cohort derived by the standardized food items.

We observed that the association of EAT-Lancet diet-adherence with stroke was stronger as compared to the association with CHD. This discrepancy may be explained by somwhat different risk profiles for CHD and stroke are comparable, but do show some differences in magnitude with low-density lipoprotein cholesterol being of particular importance for CHD, and hypertension being of greater influence for stroke ^34,35^. Additionally, several studies have found a differential effect of individual food groups with CHD and stroke. For example, high consumption of fruit, vegetables, fish, dairy (e.g., cheese, fermented dairy) and limited consumption of red and processed meat and sugar-sweetened beverages have all been related to both lower risk of stroke and CHD ^36–41^, while intakes of whole grains, nuts, and legumes have been more consistently associated with CHD rather than stroke ^36,38,40^. This may explain the inconsistent results across individual EPIC cohort studies. Furthermore, we observed that risks of both CHD and stroke associated with EAT-Lancet diet adherence were lower among those younger than 60 years old compared to the overall cohort, which may indicate that adoption of the EAT-Lancet diet is of importance for cardiovascular risk in this population.

With regards to the environmental impact of the EAT-Lancet diet, the observed lower levels of GHG emissions and land use in relation to higher adherence are modest, yet statistically significant. These modest effects may be explained by higher consumption of dairy, eggs, chicken, and fish in those with higher adherence compared to those with the least adherence. This may offset the effects on GHG emissions and land use we would expect from consuming less red and processed meat. Albeit less pronounced, the effects on environmental indicators are comparable to those we observed in the Dutch EPIC cohorts ^9^. The difference between the two studies may be related to the distinct underlying methodologies used to calculate the environmental impact of foods. This study used SHARP-ID data, which uses different databases and sources for transport than the Dutch Life Cycle Assessment (LCA) database used in Colizzi et al. ^27,42^. The two databases also considered other system boundaries, allocation methods, and geographical zones ^27,43,44^. Furthermore, SHARP-ID data does not include country-specific estimates, which may lead to different or less pronounced associations.

Of note, the impact on levels of both GHG emissions and land use was much higher when using the EAT-Lancet dietVV-score. The scoring method for meat and animal-based products are the sole difference between the two scores, underlying the impact that limiting meat and dairy products has on environmental indicators in this population. A (pro-) vegetarian or vegan version of the EAT-Lancet diet score did not particularly alter benefits to health, compared to the original score, but largely impacted the GHG emissions and land use associated with the diet. These findings are in line with the large body of evidence on the environmental footprint of the production and consumption of meat, and on the reduction in environmental impact that can be achieved by switching to more plant-based diets ^14,45,46^. Our findings using the EAT-Lancet dietVV-score are also in line with the reduction in GHG emissions and land use measured by similar studies assessing the environmental footprint of the EAT-Lancet diet ^15,47–50^, for example with a previous study in EPIC by Laine et al. (2021) that used counterfactual attributable fraction modelling for shifting from low adherence to full adherence to the diet, and showed a 50% reduction in GHG emissions and 62% lower land use ^50^. Although more modest, adherence to the EAT-Lancet dietVV-score — which resembles the most the scoring used by Laine et al. ^50^ — in our population shows similar co-benefits to the study by Laine et al. ^50^.

With regards to food biodiversity, to our knowledge, this is the first study measuring the DSR associated with the EAT-Lancet diet. Higher adherence to the EAT-Lancet diet was associated with higher plant species richness and lower animal species richness, but it did not reflect higher diversity in an individual’s total food consumption. These findings can be largely explained by the fact that the EAT-Lancet diet-score values higher intakes of plant-based foods and lower intakes of animal-based products. An individual with a high EAT-Lancet diet-score is likely to eat a varied group of plant species, and a small number (or no) animal species. Additionally, we excluded all other foods that could not be uniquely allocated to plant or animal species, which can explain the discrepensies between overall DSR and DSRPlant and DSRAnimal. Because of the importance of biodiversity for human nutrition and agricultural ecosystems, future research on sustainable diets could consider including DSR as a valuable tool for the development of environmental and food policies.

Lastly, we need to address some of this study’s strengths and limitations. Strengths of the current study include the large pan-European population with a substantial number of cases, the prospective design, the diversity of diets captured, and the use of validated food frequency questionnaire (FFQ) data. Furthermore, our scoring approach complies with various preferable features of a priori dietary indices, including the use of various index dimensions (e.g., adequacy, moderation, optimum, and ratio components), the use of metric measures as opposed to ordinal or dichotomous metrics, and the use of normative cut-off values ^12,13^. However, this study also has limitations. First, dietary data were self-reported and only assessed at baseline, possibly introducting misclassification of exposure status. Not only dietary habits in the population may have changed throughout the years, dietary data were collected almost 30 years before the publication of the recommendations by the EAT-Lancet Commission. Second, due to the multicenter design, non-fatal CHD and stroke events were ascertained through a variety of methods (e.g., linkage with disease registries, follow-up questionnaires), which may have led to non-differential misclassification of outcome status. Similarly, environmental impact assessment through SHARP-ID relied on data published across a number of years and may not entirely reflect the environmental impact of foods and beverages at the time of data collection ^27^. Third, despite adjusting for various demographic, lifestyle, and cardiometabolic risk factors, residual confounding bias cannot be ruled out, for instance, because of socio-economic differences and additional cardiometabolic risk factors unaccounted for in this study. Additionally, we did not observe complete adherence to the EAT-Lancet diet in our population, possibly resulting in smaller effects for cardiovascular and planetrary health.

Finally, we need to address that the EAT-Lancet diet represents a global diet that should be translated to national food-based dietary guidelines. In order to do so, additional dimensions relevant to a sustainable food system transition would need to be measured in context. These include, for instance, the affordability of the diet, cultural acceptance, and nutritional adequacy of the diet, all of which were not tested in this study. Two previous studies aimed to translate the generic EAT-Lancet diet recommendations to country-specific contexts and found country-specific EAT-Lancet diets to be nutritionally adequate, except for vitamin D, iodine and calcium ^51,52^. A recent modelling study measured the impact of adherence to EAT-Lancet diet recommendations on all-cause mortality compared to adherence to current food-based dietary guidelines ^53^. While it found that more ambitious recommendations for whole grains, nuts and seeds, legumes, vegetables, and processed meat were linked to greater reductions in mortality, it also showed that none of the 85 countries included in the analysis adhered to all food-based dietary guidelines ^53^. This highlights an important concern about the feasibility of transitioning to dietary patterns compliant with EAT-Lancet diet recommendations. Moreover, this study showed that the way in which adherence to the EAT-Lancet diet is being operationalised may result in different estimates of co-benefits for health and the environment, limiting comparability of findings on the EAT-Lancet diet and the use of these research studies as the base for the development of national policies.

To conclude, this prospective cohort study showed co-benefits of the EAT-Lancet diet for human cardiovascular health and environmental well-being in a large European cohort. This research showed that adherence to the EAT-Lancet diet was associated with lower risk of stroke but not of CHD, modestly lower GHG emissions and lower land use, higher plant species richness and lower animal species richness. Future research should further explore the role of animal-sourced foods, operationalization of the diet, nutritional adequacy and affordability of the EAT-Lancet diet.

## Data Availability

EPIC data and biospecimens are available for investigators who seek to answer important questions on health and disease in the context of research projects that are consistent with the legal and ethical standard practices of the International Agency for Research on Cancer (IARC), WHO, and the EPIC centres. The primary responsibility for accessing the data, obtained in the frame of the present publication, belongs to the EPIC centres that provided them. Access to EPIC data can be requested to the EPIC Steering Committee, as detailed in the EPIC-Europe Access Policy.

## Contributors

All authors contributed to reviewing and editing the writing of the manuscript, interpretation of statistics and findings, critically revised the article for content, and approved the final version. CC verified the underlying data, did the formal analysis, contributed to conceptualization, methodology and writing and reviewing of the original manuscript. JWJB and YTvdS contributed to the conceptualization, methodology, supervision, and writing and reviewing of the original manuscript. REV, KA, CCD, IH, TJK, JEL, KP, PV, EW, CA, JB, PC, JH, AH, VK, GM, OM, CMI, GN, DRP, CS, MJS, MBS, AT, and WMMV were working group members, representing a centre of EPIC and thus provided one of the following: data curation, project administration, or resources, and provided feedback and contributed to the writing of the original manuscript.

## Data sharing

EPIC data and biospecimens are available for investigators who seek to answer important questions on health and disease in the context of research projects that are consistent with the legal and ethical standard practices of the International Agency for Research on Cancer (IARC), WHO, and the EPIC centres. The primary responsibility for accessing the data, obtained in the frame of the present publication, belongs to the EPIC centres that provided them. Access to EPIC data can be requested to the EPIC Steering Committee, as detailed in the <u>EPIC-Europe Access Policy</u>.

## Sources of Funding

The coordination of EPIC-Europe is financially supported by International Agency for Research on Cancer (IARC) and also by the Department of Epidemiology and Biostatistics, School of Public Health, Imperial College London which has additional infrastructure support provided by the NIHR Imperial Biomedical Research Centre (BRC).

The national cohorts are supported by: Danish Cancer Society (Denmark); German Cancer Aid, German Cancer Research Center (DKFZ), German Institute of Human Nutrition Potsdam-Rehbruecke (DIfE), Federal Ministry of Education and Research (BMBF) (Germany); Associazione Italiana per la Ricerca sul Cancro-AIRC-Italy, Italian Ministry of Health, Italian Ministry of University and Research (MUR), Compagnia di San Paolo (Italy); Dutch Ministry of Public Health, Welfare and Sports (VWS), the Netherlands Organisation for Health Research and Development (ZonMW), World Cancer Research Fund (WCRF), (The Netherlands); Instituto de Salud Carlos III (ISCIII), Regional Governments of Andalucía, Asturias, Basque Country, Murcia and Navarra, and the Catalan Institute of Oncology - ICO (Spain); Swedish Cancer Society, Swedish Research Council and County Councils of Skåne and Västerbotten (Sweden); Cancer Research UK (C864/A14136 to EPIC-Norfolk; C8221/A29017 to EPIC-Oxford), Medical Research Council (MR/N003284/1, MC-UU_12015/1 and MC_UU_00006/1 to EPIC-Norfolk; MR/Y013662/1 to EPIC-Oxford) (United Kingdom).

Previous support has come from “Europe against Cancer” Programme of the European Commission (DG SANCO).

The funders had no role in study design, data collection and analysis, decision to publish, or preparation of this manuscript.

## Acknowledgments

We acknowledge that Hyblean Association for Epidemiology Research, AIRE ONLUS Ragusa, Italy is data contributor for EPIC-Ragusa.

## Disclaimer

Where authors are identified as personnel of the International Agency for Research on Cancer / World Health Organization, the authors alone are responsible for the views expressed in this article and they do not necessarily represent the decisions, policy or views of the International Agency for Research on Cancer / World Health Organization.

