## Supplemental Material for "The EAT-Lancet Planetary Health Diet: Impact on Cardiovascular Disease and the Environment in the EPIC Cohort"

Chiara Colizzi et al.

### **Supplemental Figures**

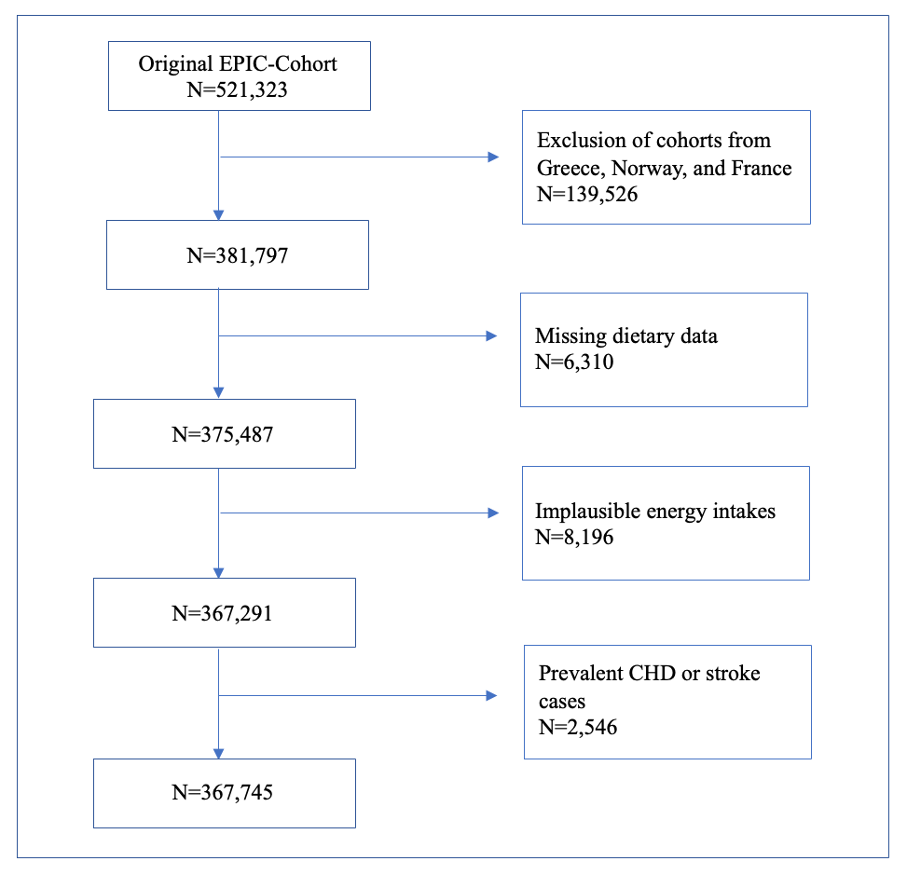
**Supplemental Figure 1. Flowchart of the study population.**

10

*Points*

*Intake (g/day)*

10

*Points*

*Intake (g/day)*

10

*Points*

*Intake (g/day)*

10

*Points*

*Ratio*

A. **Adequacy component**
B. **Moderation component**
C. **Optimum component**
D. **Ratio component**

**Supplemental Figure 2. Graphical illustration of the scoring components applied to EAT-Lancet diet-score.**

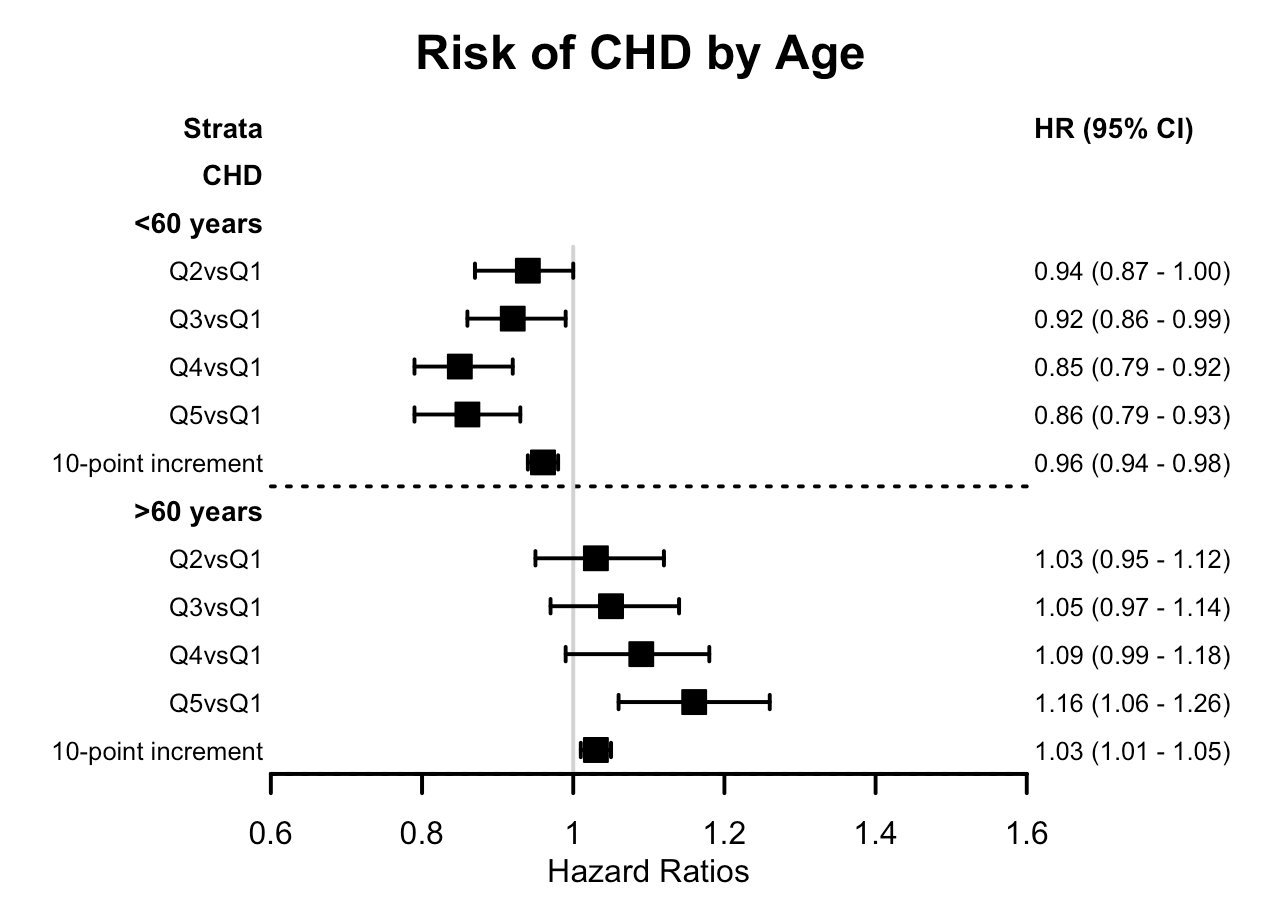

**Supplemental Figure 3. Hazard ratios and 95% confidence intervals for the association between EAT-Lancet diet score and incident CHD by age groups.^1^**

**^1^** All analyses adjusted for age, sex, educational level, smoking, alcohol consumption, physical activity, energy intake, BMI, waist circumference, hypertension, hyperlipidemia, and diabetes status, and stratified by EPIC center.

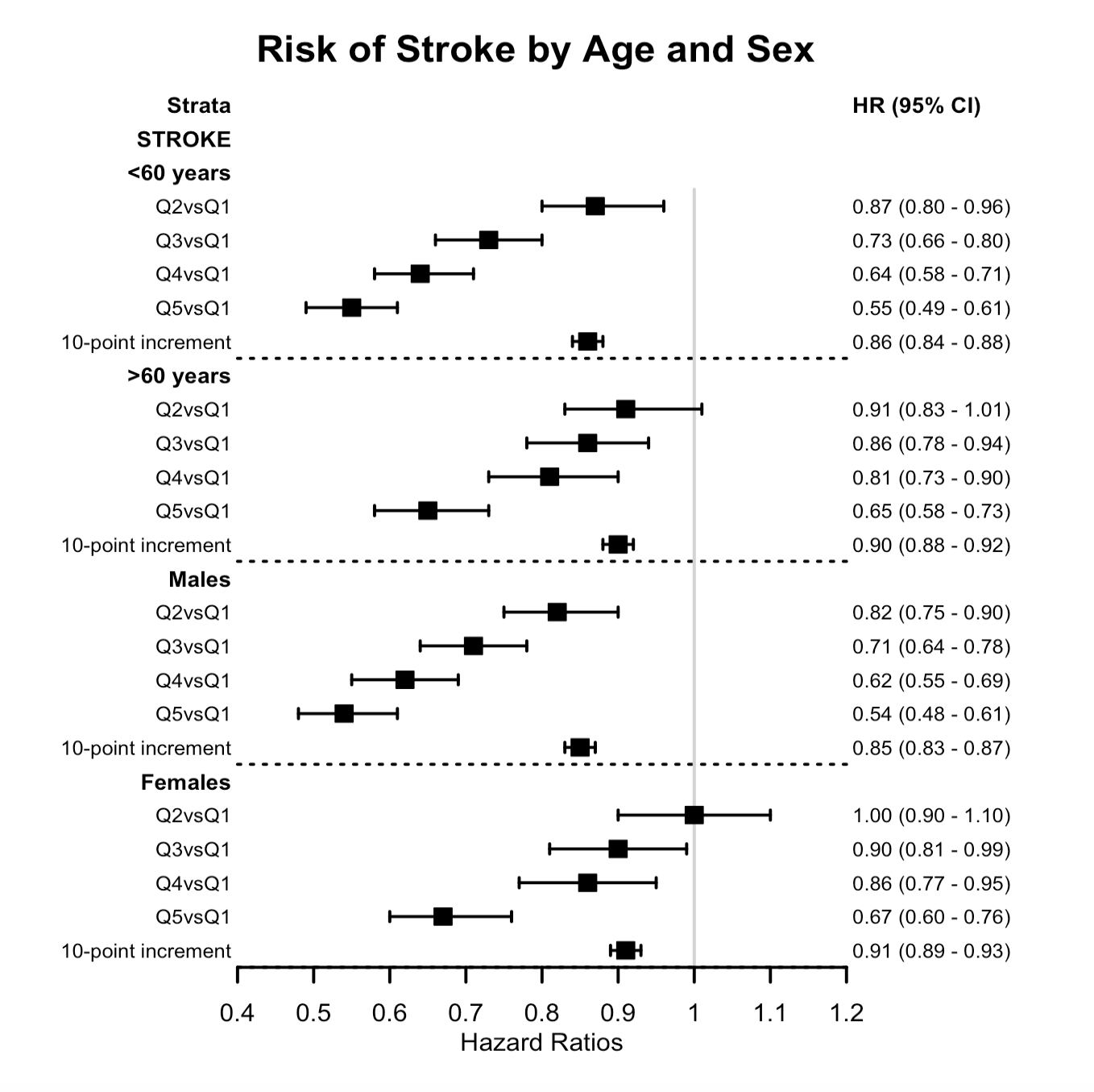

**Supplemental Figure 4. Hazard ratios and 95% confidence intervals for the association between EAT-Lancet diet score and incident stroke by age and sex.^1^**

**^1^** All analyses adjusted for age, sex, educational level, smoking, alcohol consumption, physical activity, energy intake, BMI, waist circumference, hypertension, hyperlipidemia, and diabetes status, and stratified by EPIC center.

### **Supplemental Tables**

**Supplemental Table 1. Scoring approach for the Eat-Lancet Planetary Health Diet score.^1^**

|  | **Scoring component** | **Points (0-10)** | | | | |
| --- | --- | --- | --- | --- | --- | --- |
|  |  | **0** | **5** | **10** | **5** | **0** |
| 1. Rice, wheat, corn etc.^2^ |  |  |  |  |  |  |
| a. Limit consumption | ~ | > 464 g/d | ≤ 464 g/d |  |  |  |
| b. Fiber intake | A | 0 g/d | ≥ 30 g/d |  |  |  |
| 2. Potatoes & cassava | O | 0 g/d |  | 50 ≤ g/day ≤ 100 |  | ≥ 150 g/d |
| 3. All vegetables | A | 0 g/d |  | ≥ 300 g/d |  |  |
| 4. All fruit | A | 0 g/d |  | ≥ 200 g/d |  |  |
| 5. Dairy | O | 0 g/d |  | 250 ≤ g/day ≤ 500 |  | ≥ 750 g/d |
| 6. Beef, lamb, pork^3^ | M | ≥ 28 g/d |  | < 14 g/d |  |  |
| 7. Chicken, poultry | O | 0 g/d |  | 29 ≤ g/day ≤ 58 |  | ≥ 87 g/d |
| 8. Eggs | O | 0 g/d |  | 13 ≤ g/day ≤ 25 |  | ≥ 38 g/d |
| 9. Fish | O | 0 g/d |  | 28 ≤ g/day ≤ 100 |  | ≥ 128 g/d |
| 10. Beans, lentils, peas | A | 0 g/d |  | ≥ 50 g/d |  |  |
| 11. Soy foods | A | 0 g/d |  | ≥ 25 g/d |  |  |
| 12. Peanuts/tree nuts^4^ | A | 0 g/d |  | ≥ 50 g/d |  |  |
| 13. Added fats^5^ | R | 13 |  | 0.6 |  |  |
| 14. Added sugar | M | ≥ 31 g/d |  | < 0 g/d |  |  |

^1^ Values represent intakes for men (based on 2500 kcal/d); intake values for women were re-calculated to 2000 kcal/d (e.g., conversion factor of 0.8).

^2^ The EAT-Lancet diet Commission proposed 232g of dry rice, wheat, and corn so EAT-Lancet diet reference level was converted to wet weight with a conversion factor of 2.

^3^ EAT-Lancet diet reference levels collapsed for a) beef and lamb and b) pork.

^4^EAT-Lancet diet reference levels collapsed a) tree nuts and b) peanuts.

^5^ Expressed as a ratio of unsaturated to saturated fat.

**Supplemental Table 2. Sub-scores on food components included in EAT-Lancet diet-score across countries.**

|  | **Italy** |  | **Spain** |  | **UK** |  | **NL** |  | **Germany** |  | **Sweden** |  | **Denmark** |  | **Full cohort** |  |
| --- | --- | --- | --- | --- | --- | --- | --- | --- | --- | --- | --- | --- | --- | --- | --- | --- |
| **Food group** | **Mean** | **SD** | **Mean** | **SD** | **Mean** | **SD** | **Mean** | **SD** | **Mean** | **SD** | **Mean** | **SD** | **Mean** | **SD** | **Mean** | **SD** |
| Rice, wheat, corn etc. | 7·2 | 2·0 | 8·7 | 1·2 | 8·4 | 1·5 | 8·6 | 1·1 | 8·5 | 1·0 | 8·0 | 1·3 | 8·9 | 1·1 | 8·3 | 1·4 |
| Potatoes & cassava | 5·3 | 3·1 | 6·7 | 3·8 | 5·9 | 4·2 | 5·5 | 4·3 | 6·6 | 3·9 | 3·8 | 4·3 | 3·7 | 4·3 | 5·3 | 4·2 |
| All vegetables | 6·3 | 2·7 | 7·6 | 2·7 | 8·3 | 2·2 | 5·2 | 2·0 | 4·7 | 2·1 | 4·5 | 2·9 | 6·2 | 2·8 | 6·2 | 2·9 |
| All fruit | 9·4 | 1·8 | 8·5 | 3·0 | 8·3 | 2·7 | 7·7 | 2·9 | 6·5 | 2·9 | 7·2 | 3·1 | 6·9 | 3·3 | 7·8 | 3·0 |
| Dairy | 7·0 | 3·3 | 7·3 | 3·4 | 6·4 | 3·9 | 5·9 | 4·0 | 6·8 | 3·2 | 6·5 | 3·9 | 5·9 | 3·9 | 6·5 | 3·7 |
| Beef, lamb, pork | 0·4 | 1·8 | 0·7 | 2·3 | 4·2 | 4·8 | 0·5 | 2·1 | 0·5 | 2·0 | 0·5 | 2·0 | 0·1 | 1·0 | 1·3 | 3·2 |
| Chicken, poultry | 6·8 | 3·2 | 6·6 | 3·7 | 4·2 | 3·8 | 4·2 | 3·1 | 4·1 | 3·0 | 3·3 | 3·3 | 6·0 | 3·1 | 4·9 | 3·6 |
| Eggs | 6·9 | 3·4 | 4·9 | 4·2 | 5·3 | 3·2 | 6·5 | 3·6 | 6·4 | 3·6 | 3·8 | 3·5 | 5·7 | 4·1 | 5·6 | 3·8 |
| Fish | 7·6 | 3·0 | 7·9 | 3·6 | 6·4 | 4·3 | 4·0 | 3·0 | 6·2 | 3·5 | 6·9 | 3·5 | 8·9 | 2·4 | 6·9 | 3·7 |
| Beans, lentils, peas | 2·4 | 2·9 | 7·8 | 3·0 | 4·4 | 3·4 | 2·2 | 2·1 | 1·0 | 1·3 | 0·8 | 1·8 | 0·3 | 0·5 | 2·7 | 3·4 |
| Soy foods | 0·0 | 0·0 | 0·0 | 0·5 | 2·6 | 3·8 | 0·9 | 2·1 | 0·1 | 0·2 | 0·0 | 0·5 | 0·0 | 0·0 | 0·7 | 2·2 |
| Peanuts/tree nuts | 0·2 | 0·5 | 0·9 | 2·0 | 1·3 | 2·0 | 1·9 | 2·3 | 0·8 | 1·3 | 0·2 | 0·7 | 0·4 | 0·9 | 0·8 | 1·6 |
| Added fats | 7·6 | 3·1 | 9·7 | 1·6 | 6·3 | 4·4 | 2·4 | 2·2 | 3·7 | 4·2 | 8·8 | 3·0 | 7·9 | 3·9 | 6·6 | 4·2 |
| Added sugar | 0·2 | 1·2 | 1·2 | 2·7 | 0·0 | 0·4 | 0·1 | 0·6 | 0·1 | 0·6 | 0·1 | 0·8 | 0·1 | 0·9 | 0·2 | 1·2 |
| Total EAT-Lancet diet-score | 67·3 | 12·0 | 78·5 | 11·4 | 72·1 | 12·4 | 55·3 | 11·2 | 55·8 | 11·2 | 54·4 | 12·4 | 60·9 | 11·5 | 63·9 | 14·5 |

**Supplemental Table 3. Hazard ratios and 95% confidence intervals for quintiles of EAT-Lancet diet adherence with incident CHD and stroke, excluding the first 2-years of follow-up.^1^**

|  | **Q1**  **(10-51)** | **Q2**  **(51-60)** | **Q3**  **(60-68)** | **Q4**  **(68-77)** | **Q5**  **(77-120)** | **Per 10-point increment** |
| --- | --- | --- | --- | --- | --- | --- |
| **CHD** |  |  |  |  |  |  |
| Incident cases, n | 2,523 | 2,465 | 2,383 | 2,153 | 1,918 | 11,442 |
| Person-years | 14,237 | 14,280 | 13,713 | 12,215 | 11,190 | 65,635 |
| HR (95% CI) | 1·00 | 0·97 (0·92 – 1·02) | 0·97 (0·92 – 1·03) | 0·95 (0·90 – 1·01) | 0·98 (0·93 – 1·04) | 0·99 (0·98 – 1·01) |
| **Stroke** |  |  |  |  |  |  |
| Incident cases, n | 1645 | 1532 | 1340 | 1141 | 859 | 6,517 |
| Person-years | 10,010 | 9,334 | 8,490 | 7,305 | 6,083 | 41,222 |
| HR (95% CI) | 1·00 | 0·89 (0·83 – 0·95) | 0·78 (0·73 – 0·84) | 0·72 (0·67 – 0·77) | 0·59 (0·54 – 0·64) | 0·88 (0·86 – 0·89) |

^1^Models were adjusted for age, sex, educational level, smoking, alcohol consumption, physical activity, energy intake, BMI, waist circumference, hypertension, hyperlipidemia, and diabetes status, and stratified by EPIC center.

| **Excluded EAT-Lancet diet recommendation** | **Risk of CHD**  **HR (95% CI)**  **per 10-point increment** | **Risk of stroke**  **HR (95% CI)**  **per 10-point increment** |
| --- | --- | --- |
| None | 0·99 (0·98; 1·01) | 0·88 (0·86; 0·89) |
| 1. Rice, wheat, corn, and other | 1·00 (0·98; 1·01) | 0·88 (0·87; 0·90) |
| 2. Potatoes and cassava | 1·00 (0·99; 1·01) | 0·89 (0·87; 0·90) |
| 3. Vegetables | 0·99 (0·98; 1·00) | 0·88 (0·87; 0·90) |
| 4. Fruit | 1·00 (0·98; 1·01) | 0·99 (0·99; 0·99) |
| 5. Whole milk or derivative equivalents | 1·00 (0·99; 1·01) | 0·88 (0·87; 0·90) |
| 6. Beef, lamb, and pork | 0·99 (0·98; 1·00) | 0·88 (0·87; 0·90) |
| 7. Chicken and other poultry | 0·99 (0·98; 1·00) | 0·88 (0·87; 0·90) |
| 8. Eggs | 0·99 (0·98; 1·01) | 0·89 (0·88; 0·91) |
| 9. Fish | 1·00 (0·99; 1·02) | 0·87 (0·86; 0·88) |
| 10. Dry beans, lentils, and peas | 0·98 (0·97; 0·99) | 0·88 (0·87; 0·90) |
| 11. Soy foods | 0·99 (0·98; 1·00) | 0·88 (0·87; 0·90) |
| 12. Peanuts and tree nuts | 0·99 (0·98; 1·00) | 0·89 (0·87; 0·90) |
| 13. Added fats | 1·00 (0·98; 1·01) | 0·85 (0·84; 0·87) |
| 14. Added sugars | 0·99 (0·98; 1·01) | 0·88 (0·87; 0·89) |

**Supplemental Table 4. Hazard ratios and 95% confidence intervals for the association between EAT-Lancet diet-scores (continuous) and incident CHD and stroke excluding one out of the fourteen recommendations individually.^1^**

^1^Models were adjusted for age, sex, educational level, smoking, alcohol consumption, physical activity, energy intake, BMI, waist circumference, hypertension, hyperlipidemia, and diabetes status, and stratified by EPIC center.

**Supplemental Table 5. Hazard ratios and 95% confidence intervals for quintiles of EAT-Lancet diet adherence with incident CHD, by EPIC country and centers. ^1^**

| **CHD** | **N/Incident cases** | **Q1**  **(10-51)** | **Q2**  **(51-60)** | **Q3**  **(60-68)** | **Q4**  **(68-77)** | **Q5**  **(77-120)** | **Per 10-point increment** |
| --- | --- | --- | --- | --- | --- | --- | --- |
| **HR, 95% CI** |  |  |  |  |  |  |  |
| Italy | 45,881 / 901 | 1·00 | 0·81 (0·64, 1·02) | 0·79 (0·63, 0·99) | 0·72 (0·57, 0·90) | 0·69 (0·54, 0·88) | 0·93 (0·87, 0·98) |
| Spain | 40,432 / 1,240 | 1·00 | 1·32 (0·80, 2·19) | 1·01 (0·63, 1·63) | 1·05 (0·66, 1·67) | 0·99 (0·63, 1·56) | 0·95 (0·90, 0·99) |
| United Kingdom | 80,058 / 3,724 | 1·00 | 1·04 (0·90, 1·20) | 1·05 (0·91, 1·20) | 0·98 (0·86, 1·12) | 1·00 (0·87, 1·15) | 0·99 (0·99, 1·01) |
| UK Health Conscious | 55,812 / 1,494 | 1·00 | 0·93 (0·71, 1·21) | 1·02 (0·79, 1·31) | 0·87 (0·68, 1·11) | 0·90 (0·70, 1·15) | 0·97 (0·93, 1·01) |
| UK general population | 24,246 / 2,230 | 1·00 | 1·10 (0·92, 1·30) | 1·05 (0·89, 1·24) | 1·04 (0·89, 1·23) | 1·11 (0·93, 1·31) | 1·01 (0·97, 1·05) |
| The Netherlands | 38,735 / 1,797 | 1·00 | 0·99 (0·87, 1·11) | 0·92 (0·80, 1·04) | 0·97 (0·81, 1·15) | 0·81 (0·58, 1·13) | 0·97 (0·93, 1·02) |
| Germany | 51,776 / 640 | 1·00 | 0·94 (0·77, 1·13) | 0·91 (0·73, 1·14) | 0·68 (0·49, 0·95) | 1·05 (0·63, 1·77) | 0·92 (0·85, 0·99) |
| Sweden | 52,344 / 2,415 | 1·00 | 0·91 (0·82, 1·00) | 0·94 (0·84, 1·05) | 0·90 (0·79, 1·04) | 0·82 (0·65, 1·03) | 0·97 (0·94, 1·01) |
| Umea | 24,505 / 624 | 1·00 | 0·95 (0·78, 1·15) | 0·84 (0·60, 1·17) | 1·10 (0·60, 2·06) | 1·38 (0·19, 10·26) | 1·00 (0·92, 1·09) |
| Malmo | 25,424 / 1,791 | 1·00 | 0·93 (0·82, 1·06) | 0·99 (0·87, 1·13) | 0·92 (0·79, 1·08) | 0·83 (0·65, 1·05) | 0·98 (0·94, 1·02) |
| Denmark | 55,519 / 1,973 | 1·00 | 0·99 (0·87, 1·11) | 0·93 (0·82, 1·06) | 0·82 (0·71, 0·95) | 0·98 (0·80, 1·19) | 0·96 (0·92, 1·00) |
| Arhus | 16,104 / 582 | 1·00 | 1·14 (0·91, 1·43) | 0·98 (0·77, 1·24) | 0·87 (0·68, 1·16) | 1·29 (1·05, 1·09) | 0·99 (0·92, 1·08) |
| Copenhagen | 37,442 / 1,391 | 1·00 | 0·93 (0·80, 1·07) | 0·92 (0·79, 1·07) | 0·80 (0·68, 0·95) | 0·87 (0·68, 1·10) | 0·95 (0·90, 0·99) |
| Full Cohort | 364,745 / 12,690 | 1·00 | 0·97 (0·92, 1·02) | 0·97 (0·92, 1·03) | 0·95 (0·90, 1·01) | 0·98 (0·93, 1·04) | 0·99 (0·98, 1·01) |

^1^ All analyses adjusted for age, sex, educational level, smoking, alcohol consumption, physical activity, energy intake, BMI, waist circumference, hypertension, hyperlipidemia, and diabetes status, and stratified by EPIC center.

**Supplemental Table 6. Hazard ratios and 95% confidence intervals for quintiles of EAT-Lancet diet adherence with incident stroke, by EPIC country and centers. ^1^**

| **Stroke** | **N/Incident cases** | **Q1**  **(10-51)** | **Q2**  **(51-60)** | **Q3**  **(60-68)** | **Q4**  **(68-77)** | **Q5**  **(77-120)** | **Per 10-point increment** |
| --- | --- | --- | --- | --- | --- | --- | --- |
| **HR, 95% CI** |  |  |  |  |  |  |  |
| Italy | 45,881 / 301 | 1·00 | 0·93 (0·62, 1·39) | 0·72 (0·49, 1·08) | 0·75 (0·51, 1·11) | 0·59 (0·39, 0·91) | 0·83 (0·76, 0·92) |
| Spain | 40,432 / 692 | 1·00 | 1·00 (0·55, 1·83) | 0·75 (0·43, 1·32) | 0·72 (0·42, 1·23) | 0·62 (0·36, 1·05) | 0·91 (0·85, 0·97) |
| United Kingdom | 80,058 / 1,034 | 1·00 | 1·13 (0·87, 1·47) | 0·89 (0·69, 1·15) | 0·97 (0·76, 1·25) | 0·91 (0·71, 1·17) | 0·95 (0·90, 1·00) |
| UK Health Conscious | 24,246 / 455 | 1·00 | 1·06 (0·71, 1·58) | 0·86(0·59, 1·27) | 0·92 (0·64, 1·34) | 0·87 (0·60, 1·27) | 0·94 (0·88, 1·00) |
| UK general population | 55,812 / 579 | 1·00 | 1·18 (0·83, 1·67) | 0·90 (0·64, 1·27) | 1·00 (0·72, 1·39) | 0·88(0·61, 1·26) | 0·95 (0·88, 1·03) |
| The Netherlands | 38,735 / 552 | 1·00 | 0·99 (0·81, 1·22) | 0·77 (0·60, 0·99) | 0·90 (0·66, 1·23) | 0·89 (0·51, 1·53) | 0·94 (0·86, 1·02) |
| Germany | 51,776 / 504 | 1·00 | 0·88 (0·71, 1·09) | 0·91 (0·72, 1·17) | 0·85 (0·62, 1·18) | 0·79 (0·43, 1·46) | 0·94 (0·86, 1·01) |
| Sweden | 52,344 / 2,166 | 1·00 | 0·94 (0·85, 1·05) | 0·92 (0·81, 1·03) | 0·87 (0·75, 1·00) | 0·70 (0·55, 0·90) | 0·93 (0·90, 0·97) |
| Umea | 25,129 / 630 | 1·00 | 0·97 (0·81, 1·17) | 0·75 (0·55, 1·03) | 0·99 (0·58, 1·70) | 0·78 (0·11, 5·72) | 0·93 (0·86, 1·02) |
| Malmo | 27,215 / 1536 | 1·00 | 0·99 (0·86, 1·14) | 1·01 (0·88, 1·17) | 0·93 (0·79, 1·10) | 0·76 (0·59, 0·99) | 0·95 (0·91, 1·00) |
| Denmark | 55,519 / 1,839 | 1·00 | 0·84 (0·74, 0·96) | 0·82 (0·72, 0·93) | 0·76 (0·65, 0·87) | 0·70 (0·56, 0·86) | 0·90 (0·86, 0·94) |
| Arhus | 16,686 / 592 | 1·00 | 0·93 (0·74, 1·16) | 0·88 (0·70, 1·10) | 0·75 (0·57, 0·98) | 0·71 (0·48, 1·07) | 0·89 (0·82, 0·96) |
| Copenhagen | 38,833 / 1,247 | 1·00 | 0·80 (0·69, 0·94) | 0·79 (0·68, 0·93) | 0·75 (0·63, 0·90) | 0·69 (0·53, 0·89) | 0·90 (0·86, 0·95) |
| Full Cohort | 364,745 / 7,126 | 1·00 | 0·89 (0·83, 0·95) | 0·78 (0·73, 0·84) | 0·72 (0·67, 0·77) | 0·59 (0·54, 0·64) | 0·88 (0·86, 0·89) |

^1^ All analyses adjusted for age, sex, educational level, smoking, alcohol consumption, physical activity, energy intake, BMI, waist circumference, hypertension, hyperlipidemia, and diabetes status, and stratified by EPIC center.
